# Smoking-informed methylation and expression QTLs in human brain and colocalization with smoking-associated genetic loci

**DOI:** 10.1101/2023.09.18.23295431

**Authors:** Megan Ulmer Carnes, Bryan C. Quach, Linran Zhou, Shizhong Han, Ran Tao, Meisha Mandal, Amy Deep-Soboslay, Jesse A. Marks, Grier P. Page, Brion S. Maher, Andrew E. Jaffe, Hyejung Won, Laura J. Bierut, Thomas M. Hyde, Joel E. Kleinman, Eric O. Johnson, Dana B. Hancock

**Affiliations:** Genomics and Translational Research Center, RTI International, Research Triangle Park, North Carolina; Lieber Institute for Brain Development (LIBD), Baltimore, Maryland; Fellow Program, RTI International, Research Triangle Park, North Carolina; Department of Psychiatry, Washington University in St. Louis, Missouri; Department of Psychiatry and Behavioral Sciences, Johns Hopkins University, Baltimore, Maryland; Department of Genetics, University of North Carolina at Chapel Hill, Chapel Hill, North Carolina; Department of Neurology, Johns Hopkins University, Baltimore, Maryland Corresponding Author: Dana B. Hancock, PhD

**Author notes:** Co-first authors (Megan Ulmer Carnes, Bryan C. Quach). These authors contributed equally. Corresponding Author: Dana B. Hancock, PhD.

## Abstract

Smoking is a leading cause of preventable morbidity and mortality. Smoking is heritable, and genome-wide association studies (GWAS) of smoking behaviors have identified hundreds of significant loci. Most GWAS-identified variants are noncoding with unknown neurobiological effects. We used genome-wide genotype, DNA methylation, and RNA sequencing data in postmortem human nucleus accumbens (NAc) to identify *cis*-methylation/expression quantitative trait loci (meQTLs/eQTLs), investigate variant-by-cigarette smoking interactions across the genome, and overlay QTL evidence at smoking GWAS-identified loci to evaluate their regulatory potential. Active smokers (N=52) and nonsmokers (N=171) were defined based on cotinine biomarker levels and next-of-kin reporting. We simultaneously tested variant and variant-by-smoking interaction effects on methylation and expression, separately, adjusting for biological and technical covariates and using a two-stage multiple testing approach with eigenMT and Bonferroni corrections. We found >2 million significant meQTL variants (p_adj_<0.05) corresponding to 41,695 unique CpGs. Results were largely driven by main effects; five meQTLs, mapping to *NUDT12*, *FAM53B*, *RNF39*, and *ADRA1B*, showed a significant interaction with smoking. We found 57,683 significant eQTLs for 958 unique eGenes (p_adj_<0.05) and no smoking interactions. Colocalization analyses identified loci with smoking-associated GWAS variants that overlapped meQTLs/eQTLs, suggesting that these heritable factors may influence smoking behaviors through functional effects on methylation/expression. One locus containing *MUSTIN1* and *ITIH4* colocalized across all data types (GWAS + meQTL + eQTL). In this first genome-wide meQTL map in the human NAc, the enriched overlap with smoking GWAS-identified genetic loci provides evidence that gene regulation in the brain helps explain the neurobiology of smoking behaviors.

## Introduction

Genetic variants that act as quantitative trait loci (QTLs) for gene regulatory features, such as DNA methylation (DNAm) and RNA expression (RNAexp) levels, are pervasive across the genome ^1–4^ Moreover, QTL variants are enriched among disease-associated loci.^5–7^ Genome-wide variants have been characterized for their QTL effects across different regulatory features, tissues, and intrinsic factors, such as sex, by the Genotype-Tissue Expression (GTEx) project ^1,8^ and others.^2,4^ However, understanding QTL effects in the context of exogenous exposures, especially in trait-relevant tissues, remains relatively limited, presenting an important next step to better understand mechanisms that underlie complex disease processes.

Cigarette smoking is an exogenous exposure associated with altered gene regulation, as shown before in both blood^9^ and brain.^10,11^ Although some gene regulation features and QTL variants are shared across tissues, others can be highly tissue-specific, with brain tissues showing the most distinct profiles compared with other tissues.^12^ Notably, poor correlations have been observed between RNAexp^1^ and DNAm levels,^13^ and differences in the QTLs implicated^14^ in brain versus other tissues means that more accessible tissues, such as blood, provide insufficient proxies for brain-specific regulatory mechanisms. Even within the brain, tissues can differ in their gene regulation levels and QTL variants that drive regulation.^1^ It is therefore critical to focus on disease-relevant tissues in the brain by relying on human postmortem tissues to assess the neurobiological underpinnings of disease-associated variants and their target genes and the neurobiological impact of cigarette smoking. By focusing on brain tissues, this study aims to shed light on the intricate mechanisms linking genetically driven gene regulation in the context of cigarette smoking in the brain, offering crucial insights into the broader health implications.

Postmortem human brain studies of various addiction-relevant tissues helped explain earlier genetic loci identified via genome-wide association study (GWAS) analyses for cigarette smoking behaviors, specifically *cis*-expression QTL (*cis*-eQTL) and *cis*-methylation QTL (*cis*-meQTL) variants.^15–17^ Since then, the genetic architecture of smoking has rapidly evolved with GWAS sample sizes exceeding 3 million in the GWAS and Sequencing Consortium of Alcohol and Nicotine use (GSCAN), resulting in hundreds of genome-wide significant loci identified for smoking initiation (ever vs. never), age at initiation, cigarettes per day, and cessation (current vs. former).^18,19^ The underlying neurobiology is unknown for most of these loci, underscoring the need to investigate their functional effects that may drive smoking behaviors. Other omics data in human brain can offer valuable insights into these effects.

Multiple brain regions are involved in cigarette smoking, and addiction more broadly, as part of a three-stage cycle.^20^ In this context, we specifically focus on nucleus accumbens (NAc), a core tissue of the first stage of the addiction cycle (binge/intoxication), given its integral role in cognitive processing of motivation, pleasure, reward, and reinforcement.^21^ To gain insights into potential functional effects of smoking-associated loci in NAc, we present a comprehensive study that maps genome-wide *cis*-eQTL and *cis*-meQTL variants and investigates variant × cigarette smoking interactions in the NAc of decedents from the Lieber Institute for Brain Development (LIBD). To our knowledge, this resource provides the first genome-wide meQTL map in human NAc, and the co-occurring *cis*-meQTL/eQTL maps in the same decedents enable a deeper understanding of shared causal mechanisms. Our study uniquely leverages smoking status postmortem to discern variants whose quantitative effects may vary upon smoking exposure, in contrast to previous phenotype-agnostic QTL maps in the human brain.^1,14^ Additionally, by colocalizing the QTL loci with GSCAN’s smoking-associated GWAS results, we identify target genes that may provide valuable neurobiological insights underlying cigarette smoking behaviors.

## Methods

### Human Postmortem NAc Samples

Postmortem human NAc tissues were obtained from the LIBD brain collection at autopsy, as previously described.^10,22^ Decedents with DSM-5 psychiatric or substance use disorders other than nicotine were excluded in addition to those with history or evidence of brain trauma, metastatic brain cancer, neurotic pathology, neurodegenerative diseases, HIV/AIDS, hepatitis, or other communicable diseases. Information regarding demographics, substance use history, and current smoking status were collected from a next-of-kin 36-item telephone-administered questionnaire (LIBD Autopsy Questionnaire). Cotinine biomarker measures from brain and/or blood samples were measured using a standard toxicology screen (National Medical Services Labs, Inc., Willow Grove, Pennsylvania). As in Markunas et al., smoking cases were defined by cotinine levels above 12 ng/mL in blood and 12 ng/g in brain, a threshold that differentiates between active and passive smoking,^23^ and by a next-of-kin report of current smoking.^10^ Controls (nonsmokers) were defined by cotinine levels below 12 ng/mL (blood) or 12 ng/g (brain) and a next-of-kin report of no current smoking.

### Genotype Data

The genotype data used in this study were obtained from a superset of samples that were genotyped and imputed as part of the full LIBD cohort, using previously described procedures.^22^ Briefly, samples from the full cohort were genotyped across multiple Illumina microarray platforms (see Supplemental Methods) and underwent a standard quality control (QC) protocol ^24^ to remove low-quality and low-frequency variants. Haplotypes were phased using SHAPEIT^25^ and then imputed using IMPUTE2^26^ with reference to 1000 Genomes phase 3. Imputed genotype dosages were converted to hard-call genotypes for variants with imputation posterior probabilities >0.9. Over 11 million autosomal variants were carried forward for analyses.

### DNAm and RNA-seq Data Generation and Processing

DNA and RNA were extracted from NAc samples of 239 eligible LIBD decedents, as described previously.^27,28^ DNAm was measured using an Illumina Human MethylationEPIC BeadChip. As described before,^10^ DNAm data processing and QC were conducted using the R package *minfi*,^29^ which included implementing stratified quantile normalization, correcting technical artifacts using principal components (PCs) of the negative control probe intensities, and controlling for tissue sample heterogeneity by estimating neuronal cell-type proportions^30^ using the Houseman method.^31^ DNAm β-values were calculated and used in the meQTL analyses, representing the percentage of DNAm at each CpG (ratio of methylated intensities relative to the total intensity).

Following RNA extraction protocols previously described, samples were sequenced using paired-end 100 bp reads on an Illumina HiSeq3000 at LIBD.^10,32^ For each RNA-seq sample, reads were trimmed and filtered using *Trimmomatic v0.39.*^33^ The remaining read pairs were pseudo-mapped to the GENCODE v34 (ENSEMBL release 100) comprehensive gene annotation reference using *Salmon v1.1.0*,^34^ and the full GRCh38 primary genome assembly was used as a selective alignment decoy sequence^35^ to enhance the accuracy of transcript quantification. Transcript quantifications were aggregated to the gene-level counts using *tximport v1.12.3*,^36^ resulting in 60,240 GENCODE genes.

To identify potential sample swaps, pairwise genotype correlations were calculated across samples using imputed genotypes and RNA-seq derived genotype calls. Samples were either excluded or mismatches resolved if the Pearson correlation coefficient ρ >0.8 for non-matching sample IDs or ρ <0.7 for matching sample IDs. Similar QC was conducted using the DNAm data.^10^ Samples were further excluded based on RNA-seq quality metrics (see Supplemental Methods), low RIN score, discrepancies between self-reported sex and chromosome Y gene expression, and missing genotype data. Sample-level QC removed 36 samples, resulting in a post-QC RNA-seq sample size of 203. Lowly expressed genes were then removed using the exclusion criteria of ≥90% of samples with ≤10 gene counts or ≤1 transcripts per million value. For eQTL mapping, GRCh37 human genome reference coordinates were used for all gene annotations to align with the genotype data genome build. In total, 16,274 genes were considered for eQTL mapping.^31^

Of the available 239 decedents, 201 RNA-seq samples and 220 DNAm samples, with genotype and smoking data, remained following QC, including 198 samples in both datasets (intersection) and 223 samples with either RNA-seq or DNAm data (union).

### Methylation and Expression Quantitative Trait Loci (meQTL/eQTL) Mapping

We performed *cis*-meQTL mapping using imputed genetic variants and DNAm intensity β-values of probes proximal, within 500 kilobases (kb) up or downstream, to these variants. Four different meQTL mapping regression models were fit: (1) a “baseline” model to test for association between genetic variants and DNAm β-values across both smoking cases and controls; this model included age at death, sex, estimated non-neuronal cell-type proportion, PC1 for DNAm array negative control probes, PC1 for imputed genotypes, and PC2 for imputed genotypes as covariates (see Supplemental Methods for model selection details); (2) a smoking cases–only model similar to the baseline model; (3) a smoking controls–only model similar to the baseline model; and (4) an interaction model to test for associations of a genetic variant-by-smoking status interaction with DNAm β-values. All models used rank-inverse normal transformed (RINT) DNAm β-values. The smoking cases–only and controls–only models are needed to generate summary statistics used to conduct stratified two degrees-of-freedom (2DF) tests. The stratified 2DF test jointly tests for genetic variant main effects and genetic variant-by-smoking status interaction effects.^37^ Additional details of each model are provided in the Supplemental Methods.

A similar framework to meQTL mapping was applied for *cis-*expression quantitative trait loci (eQTL) mapping. Baseline, smoking cases–only, smoking controls–only, and interaction eQTL models were fit for each imputed genetic variant and NAc expression levels of genes proximal (within 500 kb of gene body) to these variants. Gene expression was represented as count values for a given gene after median-of-ratios normalization^38^ and RINT. The covariates included in each model were age at death, sex, exon mapping rate, ribosomal RNA mapping rate, PC1 for imputed genotypes, PC2 for imputed genotypes, and four latent variables estimated by PEER v1.3^39^ to account for additional unmeasured sources of confounding (see Supplemental Methods for model selection details).

To account for multiple hypothesis testing in the meQTL/eQTL models and stratified 2DF tests, p-values were corrected using a conversative two-stage approach to mitigate inflation associated with single-stage approaches when applied to QTL mapping.^40^ This hierarchical approach first accounts for association tests across variants for a given DNAm probe/gene, then accounts for tests across all probes/genes. In the first stage, all nominal p-values for a given probe/gene were adjusted using EigenMT.^41^ In the second stage, EigenMT adjusted p-values were further corrected by the number of probes/genes tested. Any initial two-stage adjusted p-value smaller than a Bonferroni corrected p-value was assigned the latter p-value as the final two-stage adjusted p-value. This ensures that the two-stage adjustment stringency does not exceed the family-wise error rate control of Bonferroni correction. A two-stage adjusted p-value cutoff of 0.05 was used to identify statistically significant QTLs.

### Gene Set Enrichment Analysis (GSEA) for eGenes and meQTL CpGs

We used the GSEAPreranked tool from GSEA v4.3.2^42^ to test for enrichment of smoking-associated eGenes and meQTL CpGs in MSigDB v2023.1.Hs gene set collections *C2 canonical pathways* and *C5 Gene Ontology gene sets*. Only gene sets with 15–500 genes were included. CpGs were assigned to genes using the Infinium MethylationEPIC v1.0 B5 Manifest, resulting in 11,772 CpGs from the interaction meQTL mapping model with at least one assigned gene. Each gene was ranked using the largest -log_10_-transformed, smoking interaction p-value for a given gene from QTL mapping. Weighted Kolmogorov-Smirnov–like statistics were computed for the enrichment scores, and p-values were determined using 1000 gene set permutations. Significant pathways were selected based on an FDR threshold of 0.10.

### QTL enrichment testing for GSCAN-identified genetic variants

We conducted variant-based enrichment testing to assess whether genome-wide significant GSCAN loci were enriched for meQTLs or eQTLs. We obtained GSCAN summary statistics from the University of Minnesota’s Data Repository for U of M (https://doi.org/10.13020/3b1n-ff32), focusing on GSCAN’s GWAS results from 2019 to capture genetic loci with variants that commonly occur and have the largest effect sizes on smoking: N up to 1.2 million individuals, depending on the smoking trait analyzed.^18^ For meQTL enrichment analysis, we compared the p-value distributions from stratified 2DF tests (i.e., stratified 2DF meQTL mapping) between GSCAN variants and a set of randomly matched variants. The random matched variant set was designed to be 10 times the size of the GSCAN variant set. The GSCAN variant set included linkage disequilibrium (LD)-pruned, genome-wide significant variants reported by GSCAN, across all four smoking traits, that were also available in our meQTL mapping (361 variants, each representing an independent significant locus). The matched variant set included 3,600 LD-pruned variants selected using SNPsnap.^43^ For variants with multiple stratified 2DF p-values (i.e., proximal to multiple CpG sites), only the smallest p-value was retained. The meQTL stratified 2DF p-value distributions for the GSCAN and matched variant sets were tested for equality using a two-sided Kolmogorov-Smirnov test. The eQTL enrichment analysis followed the same procedure as the meQTL enrichment analysis. The GSCAN variant set included 305 variants because the overlap with variants from eQTL mapping differed from meQTL mapping. The SNPsnap constructed, matched variant set included 3,050 (305 × 10) variants.

### Colocalization between GSCAN smoking GWAS and meQTL/eQTL mappings

We tested whether meQTL or eQTL signals from the baseline model QTL mapping colocalized with GSCAN loci for smoking initiation, age at initiation, cigarettes per day, and cessation using the *coloc v5.1.0* R package. For simplicity, we describe this analysis in relation to a single GSCAN trait and meQTLs, but an equivalent framework was applied for GSCAN-eQTL colocalization testing. All DNAm probes with an meQTL mapping *cis*-window that overlapped with a GSCAN locus were considered for colocalization. For a given probe, the colocalization test region spanned all genetic variants that were 1) included in the meQTL mapping for the probe and 2) tested in the GSCAN GWAS (Figure S1). Summary statistics from GSCAN and the baseline model meQTL mapping for these variants were provided to the *coloc.abf* function to perform colocalization testing. A colocalization test region was considered as having a colocalized signal between the GSCAN GWAS and meQTL mapping if the *coloc* posterior probability of hypothesis 4 (both traits are associated and share a single causal variant) exceeded 0.8 and the meQTL had a genome-wide significant, two-stage, adjusted p-value in the baseline meQTL analysis.

For GSCAN loci that showed colocalization with both an meQTL and an eQTL, HyPrColoc (https://github.com/jrs95/hyprcoloc/; commit ID f279ceb) was applied to assess whether these colocalizations resulted from the same region of the locus.^44^ Each HyPrColoc test only included one GSCAN trait, eGene, and CpG, so if multiple genes or CpGs independently colocalized with the GSCAN trait, all combinations of gene–CpG pairs were combined with the GSCAN trait for a HyPrColoc test. For each HyPrColoc test, a genetic variant was included as input only if it had summary statistics available from the baseline model eQTL/meQTL mapping and GSCAN GWAS for the smoking trait. Significant colocalization was defined as a GSCAN trait–eGene–CpG triplet having posterior probability >0.8.

## Results

### Overview

Of the available 239 decedents, 223 (52 cases and 171 controls) samples with genotype and smoking data and either RNA-seq or DNAm in NAc remained after QC (Table 1). Of the cases, 50% had African ancestry (AA) and 50% had European ancestry (EA) based on next-of-kin report and genotype confirmation. The manner of death differed slightly among cases and controls; more controls died by accident (17%) compared to cases (3.8%), and cases were more likely to die from natural causes (88.5%) compared to controls (74.9%). Age, sex, and postmortem interval were similar for cases and controls.

**Table 1.**
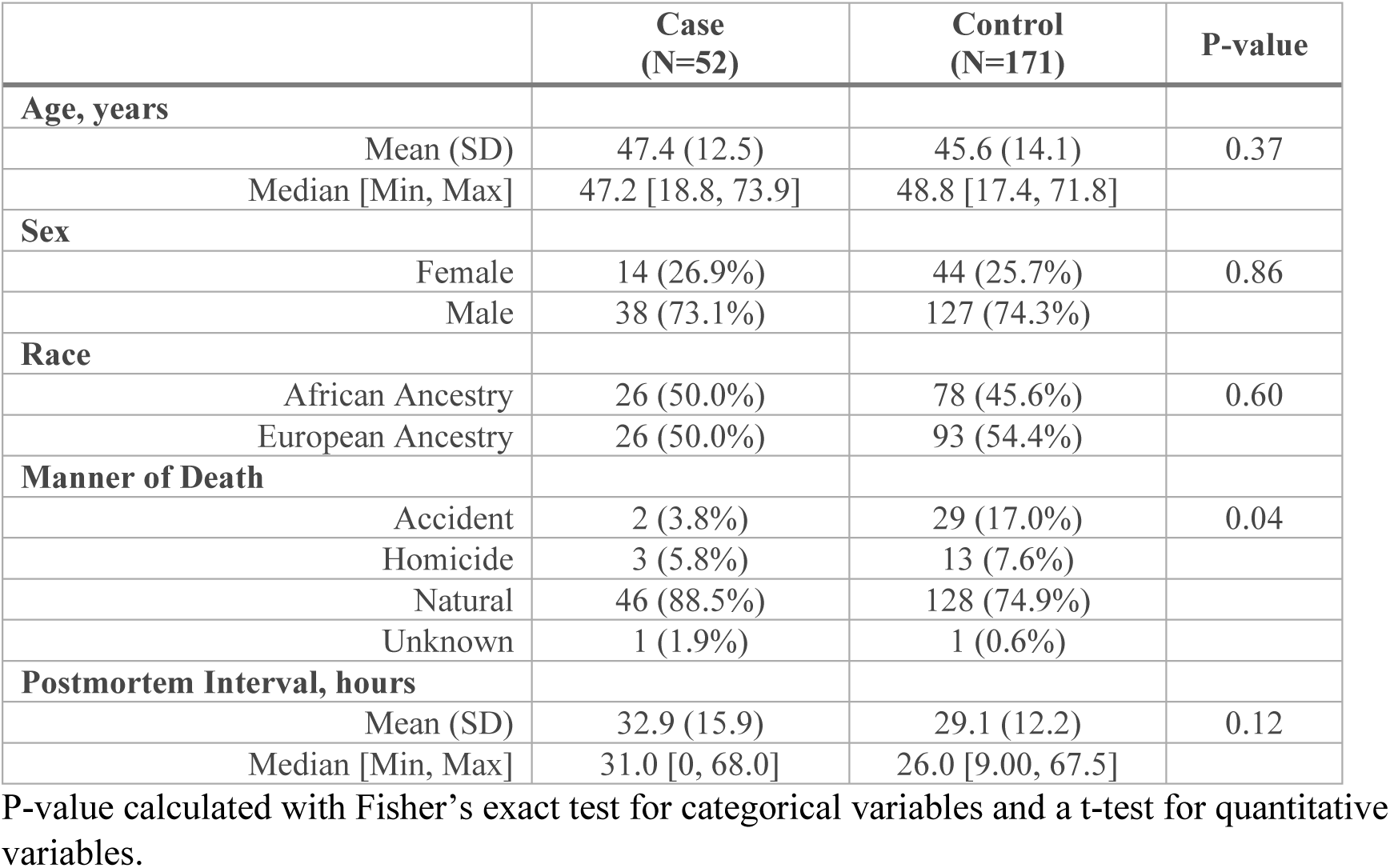
Characteristics of decedents included for methylation and expression quantitative trait loci mapping in nucleus accumbens.

|  | **Case**<br>**(N=52)** | **Control**<br>**(N=171)** | **P-value** |
| --- | --- | --- | --- |
| **Age, years** |  |  |  |
| Mean (SD) | 47.4 (12.5) | 45.6 (14.1) | 0.37 |
| Median [Min, Max] | 47.2 [18.8, 73.9] | 48.8 [17.4, 71.8] |  |
| **Sex** |  |  |  |
| Female | 14 (26.9%) | 44 (25.7%) | 0.86 |
| Male | 38 (73.1%) | 127 (74.3%) |  |
| **Race** |  |  |  |
| African Ancestry | 26 (50.0%) | 78 (45.6%) | 0.60 |
| European Ancestry | 26 (50.0%) | 93 (54.4%) |  |
| **Manner of Death** |  |  |  |
| Accident | 2 (3.8%) | 29 (17.0%) | 0.04 |
| Homicide | 3 (5.8%) | 13 (7.6%) |  |
| Natural | 46 (88.5%) | 128 (74.9%) |  |
| Unknown | 1 (1.9%) | 1 (0.6%) |  |
| **Postmortem Interval, hours** |  |  |  |
| Mean (SD) | 32.9 (15.9) | 29.1 (12.2) | 0.12 |
| Median [Min, Max] | 31.0 [0, 68.0] | 26.0 [9.00, 67.5] |  |

P-value calculated with Fisher's exact test for categorical variables and a t-test for quantitative variables.

We first generated single data type QTL (eQTL and meQTL) maps in a mega-analysis of EA and AA decedents. Significant QTLs were annotated and characterized as a main effect or as showing evidence of a significant smoking interaction. Each QTL type independently underwent colocalization analysis with GSCAN GWAS summary statistics, and a joint colocalization analyses across data types was performed for significant me/eQTL. See Figure 1 for an overview of the analysis workflow.

**Figure 1:**
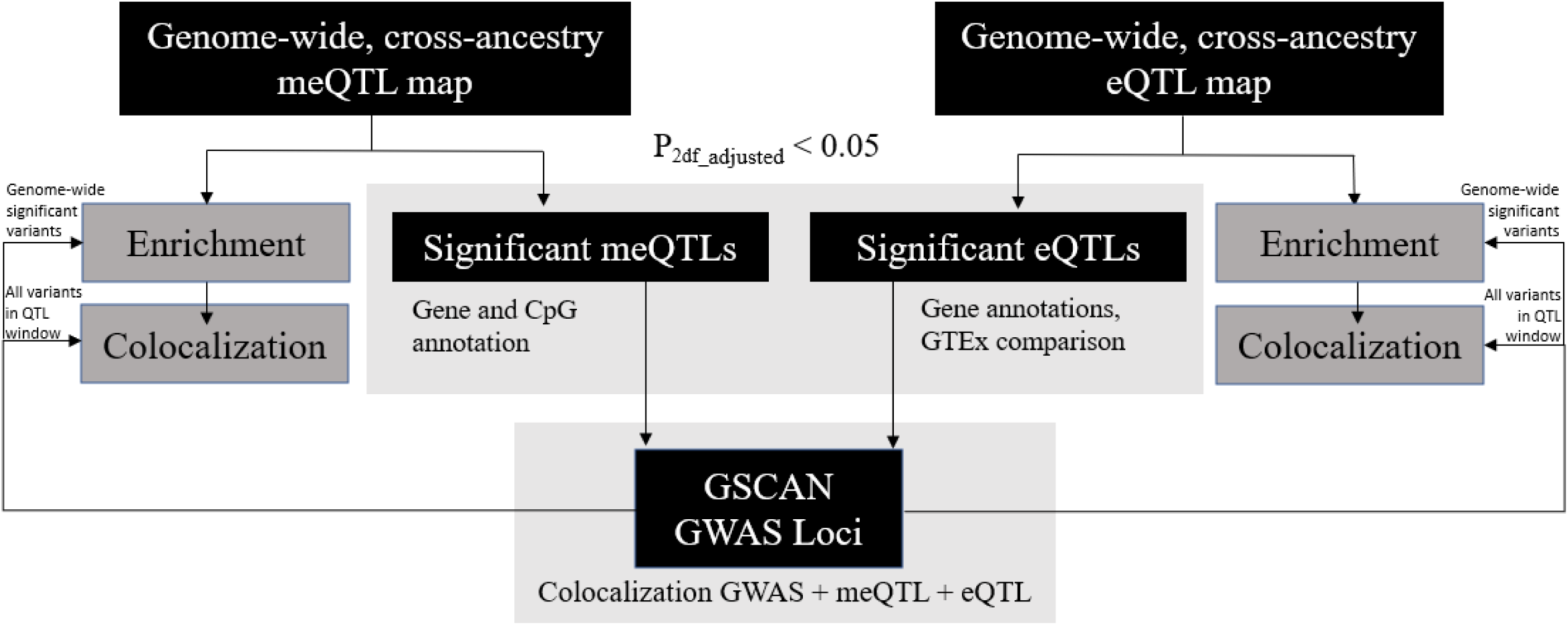
Analysis workflow. Genome-wide *cis*-eQTL and *cis*-meQTL maps were constructed using DNA methylation, gene expression, and genotype data generated on decedents in the Lieber Institute for Brain Development (LIBD) Human Brain Repository. Significant QTLs following multiple testing correction underwent further annotation. QTL maps were integrated with GSCAN GWAS summary statistics to perform genetic variant enrichment testing and colocalization analyses.

### Genome-wide cis-meQTL Maps

DNAm in NAc and genotype data were available for 52 smoking cases (26 EA, 26 AA) and 168 smoking controls (75 EA, 93 AA). A total of 11,206,899 variants were used in the initial analysis and 784,843 CpGs, resulting in 1,748,985,510 meQTL tests. After applying two-stage multiple testing correction, we identified 2,552,641 significant meQTL variants targeting 51,315 unique CpGs. These results are shown in Supplemental Table S1, which is restricted to the lead variant for each unique CpG because of the size of the meQTL analysis (Table S1, Column: P_interaction_). Full results for all variants are publicly available at synapse.org (https://doi.org/10.7303/syn50996324).

To identify the most robust signals, we performed post hoc filtering, keeping only meQTLs where the top variant had a minor allele frequency (MAF) ≥ 0.05 and missingness ≤ 0.10 in both ancestries, leaving 41,695 unique CpGs. The top five most significant meQTLs include CpGs that map to *PITRM1*, *KDM3B*, *ARID1B*, or *MTL5* (Table S1).

### Genome-wide cis-eQTL Maps

Gene expression (RNA-seq) in NAc and genotype data were available for 47 smoking cases (24 EA, 23 AA) and 156 smoking controls (72 EA, 84 AA). These data were previously used to map eQTLs agnostic to the smoking phenotype (i.e., without accounting for the main effect of smoking or variant-by-smoking interaction).^45^ In the present study, accounting for variant-by-smoking interaction and after multiple testing correction, we identified 83,095 significant eQTL variants from 1,050 eGenes. Of these, 57,683 eQTL variants targeting 958 unique eGenes remained after post hoc filtering was applied to identify the most robust signals, keeping only eQTLs where the top variant had an MAF ≥0.05 and missingness ≤0.10 in both ancestries (Table S2). All significant results are shown in Table S2. Table S3 is filtered to the lead variant for each significant eGene (N=1,050). The 10 most significant eGenes were *RPL9*, *ZSWIM7*, *GATD3B*, *RPS28*, *XRRA1*, *TMEM161B-AS1*, *CUTALP*, *NIPBL-DT*, *ZNF718*, and *SPATA7* (Table S3). Full results for all tested variants are publicly available at synapse.org (https://doi.org/10.7303/syn50996324).

Both meQTLs and eQTLs were pervasive throughout the genome (Figure 2). When comparing the eQTL to meQTL results, 562/1,050 (54%) significant eGenes overlapped genes annotated to significant meQTL CpGs. When comparing our eQTL results to GTEx, 655 of the 1,050 unique and significant eGenes identified in our analysis are present in the GTEx NAc analysis after MAF and missingness filtering. Of these 655, 509 (78%) met a Bonferroni correction significance threshold (0.05/655=7.6e-5) (Table S3, GTEx column: *Nominal p-value*). Thus, we observed a high correlation when comparing NAc eQTL results between our study and GTEx.

**Figure 2:**
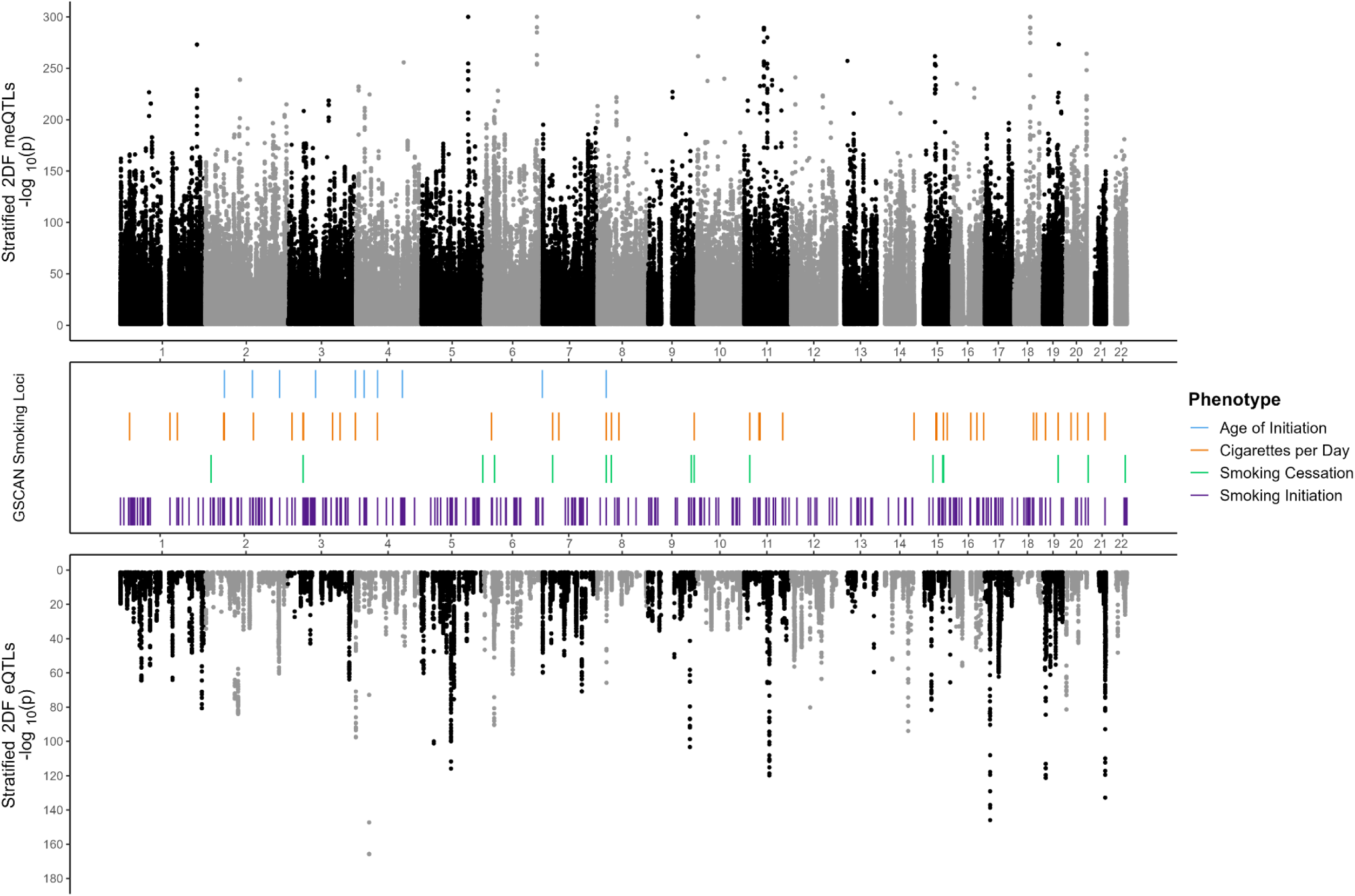
Miami plot for stratified 2DF meQTL and eQTL maps. Stratified 2DF QTL mapping -log_10_ nominal p-values for significant (2df adjusted p <= 0.05) genetic variant–CpG probe (top panel) and genetic variant-gene associations tests (bottom panel) are displayed as a function of genome position (x-axis). Genomic locations of significant GSCAN GWAS loci for four smoking phenotypes are denoted by the bars in the middle panel.

### Smoking Interaction Effects with meQTLs/eQTLs

To identify meQTLs/eQTLs that differed by smoking, we compared the primary analysis results, generated using a 2DF test, with the baseline models and interaction models. The meQTLs were primarily driven by main effects, with few showing evidence for interaction. Of the significant 41,695 unique CpGs, only five demonstrated strong evidence of an interaction with smoking (Table 2, Figure S2) based on Bonferroni correction (significance threshold of 0.05/41,695=1.2e-6). No eQTLs showed evidence of an interaction (interaction significance threshold of 0.05/877 unique eGenes=5.7e-5) after filtering by MAF and missingness.

**Table 2.** Five meQTLs with evidence of significant variant × smoking interactions.

| Variant | CpG | CpG distance from top variant (bp) | Gene | P<sub>2DF Adjusted</sub> | P<sub>main effect</sub> | P<sub>interaction</sub> |
| --- | --- | --- | --- | --- | --- | --- |
| rs16965753 | cg05215806 | 87468 | — | 0.005 | 0.76 | 2.5E-07 |
| rs73190041 | cg06105925 | 43102 | *NUDT12* | 0.002 | 1.3E-06 | 1.5E-07 |
| rs4962681 | cg17990900 | 5083 | *FAM53B* | 1.02E-08 | 2.9E-06 | 5.8E-07 |
| rs7770811 | cg20119745 | 78880 | *RNF39* | 0.045 | 2.7E-05 | 6.4E-07 |
| rs6884129 | cg25315648 | 104 | *ADRA1B* | 3.41E-20 | 1.36E-17 | 8.7E-07 |

See Table S1 for full annotations.

Because use of stringent significance thresholds may miss subtle smoking interactions with individual variants, we performed GSEA using genes ranked based on QTL–smoking interaction p-values to identify QTL-enriched biological processes and pathways that may be altered by smoking. For meQTL-smoking interactions, five pathways related to the synaptic cleft (the gap between pre-and postsynaptic membranes where neurotransmitters are released) were implicated. For eQTL-smoking interactions, cell cycle and wound response processes (cellular changes resulting from a stimulus indicated damage to an organism) were enriched (Table S4).

### GWAS-identified Variants that Exert QTL Effects on their Target Genes

GSCAN’s GWAS analyses identified 462 independent variants at 325 loci that showed significant association with at least one of four smoking phenotypes (initiation, age at initiation, cigarettes per day, and cessation)^18^. Of these variants, 361 were available in our meQTL map and 305 were available in our eQTL map. Annotating the GWAS variants to our QTL maps revealed that 63% of the GWAS-identified variants were significant meQTLs, whereas only 7% were significant eQTLs (Tables S5–S7). Only a few (eQTL N=11, meQTL N=293) were significant at the genome-wide level (Tables S5–S6, column *Two-stage adjusted p-value_2DF_*).

The GWAS-identified variants mapped to 1,201 significant variant–CpG pairs from the meQTL 2DF model, of which 135 showed nominal evidence for variant-by-smoking interaction (P_interaction_<0.05) (Table S5). Additionally, the GWAS-identified variants mapped to 27 significant variant–eGene pairs from the eQTL 2DF model, of which three showed nominal evidence for interaction (P_interaction_<0.05) (Table S6). For example, rs8192726 (located in a well-known nicotine metabolizing gene: *CYP2A6*) was associated with cigarettes per day and methylation at cg18820595 (*LTBP4*) among smokers (P_cases_=8.2×10^−6^) but not among nonsmokers (P_controls_=0.34). However, nearly all meQTL and eQTL patterns observed for GSCAN-implicated variants were driven by main effects without differences observed between smokers and nonsmokers. One variant–CpG pair overlapped with multiple smoking phenotypes: rs11780471-cg00421144, with the major allele (A) being associated with increased risk for smoking initiation, lower age at initiation, and decreased methylation level in *CHRNA2*.

To test the overlapping evidence of GWAS-identified variants as QTLs more formally, we performed an enrichment analysis using a two-sample Kolmogorov-Smirnov test that compared NAc meQTL and eQTL p-value distributions at GSCAN significant variants with a random set of variants. We found that GSCAN variants were significantly enriched for meQTLs (p-value=0.005) but not for eQTLs (p-value=0.3). For comparison, using the same test with NAc eQTLs from GTEx, we found an enrichment p-value=0.07.

### Smoking meQTL, eQTL, and GWAS Colocalization

We performed colocalization analyses of GSCAN’s GWAS results with our QTL maps to characterize heritable components of smoking that exert QTL effects. First, we performed pair-wise analyses (meQTL + GWAS, eQTL + GWAS), starting with GSCAN significant loci. Because a single locus may include more than one unique CpG or eGene, many colocalization analyses were performed per region. See Methods for additional details. In general, we observed more colocalization of meQTLs than eQTLs with the GWAS loci (Table 3, Tables S8–S9). No eQTLs that colocalized with a GSCAN locus overlapped more than one phenotype. However, four genome-wide significant CpGs colocalized across two phenotypes (cigarettes per day and age of initiation): cg12293539 at *MAML3* and cg00622170, cg11254171, and cg18236429 at the *NOP14/NOP14-AS1* locus. Additionally, smoking initiation had the most colocalizing loci, in line with being the phenotype with the most significant number of GSCAN associations.

**Table 3.** Summary of the meQTL and eQTL colocalization with the GSCAN smoking GWAS results.

| Data type overlap | Phenotype | N unique<br>eGene/CpG<br>PP(H4)>0.8 | N GSCAN unique<br>loci that colocalized /<br>total |
| --- | --- | --- | --- |
| **GSCAN-meQTL** | Smoking Initiation | 83 | 44/256 (17%) |
|  | Age of Smoking Initiation | 8 | 4/10 (40%) |
|  | Cigarettes per Day | 42 | 12/39 (31%) |
|  | Smoking Cessation | 13 | 5/16 (31%) |
| **GSCAN-eQTL** | Smoking Initiation | 3 | 2/248 (1%) |
|  | Age of Smoking Initiation | 0 | 0/9 (0%) |
|  | Cigarettes per Day | 1 | 1/39 (3%) |
|  | Smoking Cessation | 0 | 0/16 (0%) |

Full colocalization results are provided in Tables S8–S9. Table 3 summarizes meQTL/eQTL loci that surpass genome-wide statistical significance and colocalize with GSCAN smoking-associated loci. *N unique eGene/CpG PP(H4)>0.8* (column 3) corresponds to the number of eGenes or CpGs with a QTL that shows evidence of colocalization with a GSCAN locus. *N GSCAN unique loci that colocalized / total* (column 4) is the fraction of unique GSCAN loci that showed evidence of colocalization.

Next, we performed colocalization using a method (HyPrColoc) that can incorporate multiple traits (i.e., methylation, gene expression, and smoking traits) in a single analysis. Focusing on GSCAN GWAS loci that colocalized with either an eQTL or meQTL, we confirmed the GWAS–eQTL–meQTL colocalizing region at the GSCAN smoking initiation locus chr3: 52386605–54266212. In total, three variant–CpG–eGene combinations had a ≥80% probability of colocalization, all involving ENSG00000243696 (predicted read-through of *MUSTN1-ITIH4*) as the eGene with (1) rs6445538 and cg25643088, (2) rs6445538 and cg19713033, and (3) rs4687472 and cg23815702. The colocalizing region encompassed a single locus on chromosome 3 containing *MUSTN1*, *STIMATE*, and *ITIH4*, in addition to other genes (Figure 3).

**Figure 3:**
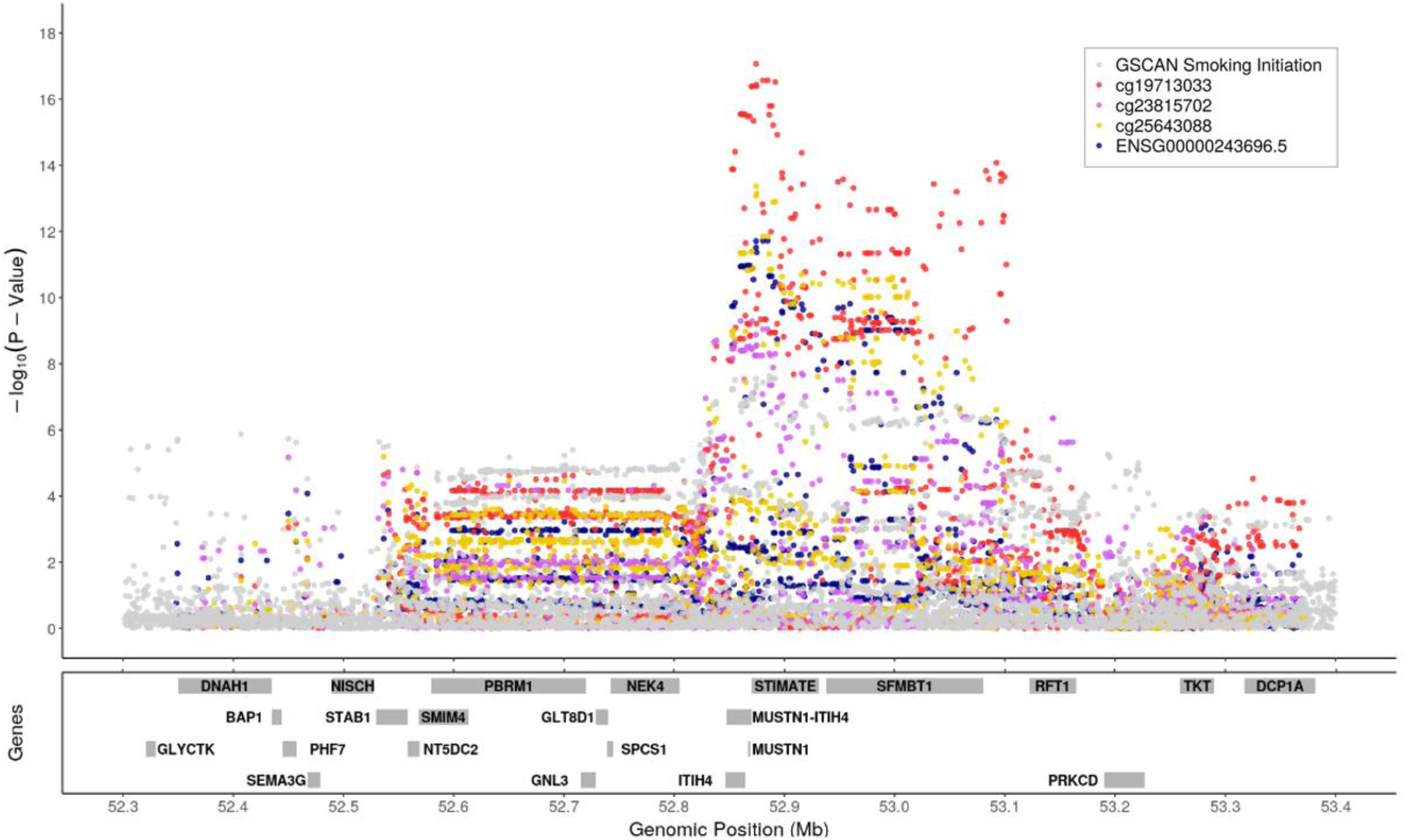
GSCAN smoking initiation GWAS, meQTL, and eQTL association test -log10 p-values near GSCAN locus chr3:52386605-54266212. -log_10_ nominal p-values for associations between genetic variants and smoking initiation, expression of three CpGs, and gene expression for the eGene identified by HyPrColoc with colocalization probability >0.8. Only genetic variants that overlapped across summary statistics for GSCAN, meQTL mapping, and eQTL mapping are plotted. Locations of genes included in the eQTL mapping are annotated in the bottom panel.

## Discussion

Cigarette smoking remains highly prevalent and a leading cause of death globally, despite decades of research into the health consequences and public health campaigns to curb smoking.^46,47^ Addiction to cigarette smoking is a complex, multi-stage process involving a neuronal rewards system that includes the NAc region of the brain.^20,48^ The NAc is known to have a role in cognitive processing of motivation, reward, and reinforcement, which are essential to the first stage of addiction (binge/intoxication).^21^ To better understand the functional effects of heritable factors that influence smoking behaviors, this study focused on identifying, for the first time, meQTLs in the NAc; investigating genetic variant interactions with cigarette smoking for both eQTLs and meQTLs; and comparing the generated QTL maps with GWAS results to better understand the neurobiology of smoking-related traits.

QTLs are pervasive throughout the genome and provide valuable insight into a tissue’s biology and gene regulation. Some QTLs are shared across tissues, whereas others exhibit tissue specificity. The GTEx project, widely used for exploring eQTLs, recently released a large-scale meQTL dataset encompassing nine human tissues.^49^ However, this dataset did not capture brain tissues. To our knowledge, ours is the first genome-wide meQTL map in human NAc. This genome-wide meQTL map, together with the genome-wide eQTL map from an overlapping dataset of the same decedents, has the potential to provide multi-omics insight into regulatory mechanisms related to many human diseases and traits. These maps are provided as a new resource to the scientific community.

Overall, we found few significant variant-by-smoking interaction effects and conclude that most QTLs in the NAc may not differ by smoking. While some interactions with small effect sizes may exist, they may require larger sample sizes to detect. We investigated this possibility using a pathway analysis ranked by evidence of smoking interactions, and we identified pathways related to cellular damage, cell cycle, and the synaptic cleft, where the nicotinic acetylcholine receptors play an important role in regulating neurotransmission.^50^ Five individual meQTLs differed significantly between smokers and nonsmokers, including Nudix Hydrolase 12 (*NUDT12*), Family with sequence similarity 53 member B (*FAM53B*), and Ring Finger Protein 39 (*RNF39*). *NUDT12* plays a role in nicotinate and nicotinamide metabolism.^51^ Interestingly, *NUDT12* was identified in a transcriptome analysis of neurons following chronic nicotine exposure ^52^ and lies within a QTL interval for nicotine sensitivity in mouse studies,^53^ as annotated in GeneWeaver.^54^ Another gene, *FAM53B*, has been associated with cocaine dependence.^55^ Differential DNAm at *FAM53B* has been observed in COPD cases compared with controls,^56^ suggesting alterations may occur in the presence of a smoking exposure. Likewise, *RNF39* was found to be differentially methylated in a study of marijuana use ^57^ and other studies of smoking-related DNAm changes.^58,59^ These findings add evidence for a select few biologically plausible genes whereby smoking may alter genetically driven gene regulation in NAc; additional evidence will require larger sample sizes to detect more subtle smoking-related effects.

To understand the shared causal variants between heritable factors (smoking-associated variants) and meQTLs/eQTLs, we employed colocalization analyses, which have not been applied with these data previously.^45^ Genomic regions with variants showing evidence of colocalized signals from both GWAS and QTL mapping suggest that the trait-associated variants act as regulators of DNAm or gene expression. We generally observed more overlap between the GWAS-associated variants and meQTLs than eQTLs; this pattern was also supported by the enrichment analyses. This observation may relate to more meQTL tests overall. However, the same pattern holds when looking at only QTLs that survive genome-wide multiple testing correction, whereby declaring meQTLs as statistically significant was based on a more stringent threshold than eQTLs and is consistent with studies of the prefrontal cortex,^60^ blood,^61^ and other tissues.^49^

One genomic region showed robust evidence of colocalization with all three data types; highlighting novel functional evidence where changes to DNAm and gene expression may help explain the neurobiology underlying a smoking initiation associated heritable factor. The primary gene indicated is a predicted read-through of the Musculoskeletal, Embryonic Nuclear Protein 1 (*MUSTN1*) and inter-alpha-trypsin inhibitor, heavy chain 4 (*ITIH4*) genes. Little is known about the read-through transcript. *MUSTN1* is known to have a role in skeletal muscle homeostasis, chondrocyte differentiation, and limb morphogenesis.^62^ *MUSTN1* expression in the brain has been observed at low levels.^1,63,64^ Schizophrenia-associated variants in *ITIH4* have been shown to regulate expression of *ITIH4* in prefrontal cortex,^65^ and ITIH4 is a biomarker for COPD.^66,67^ *ITIH4* is also expressed at low levels in the brain and is largely expressed in the liver, although it is also moderately expressed in skeletal muscles like *MUSTN1.*^1,63,64^ Interestingly, there is increasing evidence that changes to skeletal muscle homeostasis can influence the physiology of the brain.^68,69^

This study has limitations to consider in interpreting the findings. First, because this study utilized an understudied brain tissue collected from decedents with a unique set of multi-omics data types (DNAm, RNA-seq, and genotypes) and smoking status, sample availability was limited to 52 smoking cases and 171 smoking controls. This constrained sample size may have limited our ability to identify interaction effects, and it limited interrogation of ancestry-or sex-specific effects and extension into independent replication datasets.

Second, QTLs were tested using a *cis*-window that still resulted in a large number of total tests, increasing the chance of type I error. We accounted for multiple testing using a two-stage correction strategy designed for this type of study^41^; however, the possibility of type I error remains. *trans*-QTL effects also merit future research with a larger sample size.

Finally, the QTL–GWAS colocalization was based on the first GSCAN meta-analysis, including up to 1.2 million individuals.^18^ GSCAN recently released an updated meta-analysis with >3 million individuals, enabling more statistical power to identify genetic loci associated with smoking traits, particularly loci with lower MAF and effect sizes than those observed in the first GSCAN meta-analysis. Although comparison with the updated GSCAN meta-analysis might result in the identification of additional QTL–GWAS colocalization signals, given the sample size available with multi-omics data in our postmortem human brain study, we would have less statistical power to detect colocalization with the lower MAF variants and smaller effect sizes observed from the additional loci identified in GSCAN2. The present study offers a comprehensive capture of a large number of genetic loci with common variants with the largest effect sizes on smoking.

Additionally, this study had several strengths. It is the first to provide a genome-wide meQTL map in human NAc, a relatively understudied brain tissue with an important role in the addiction cycle. Case definitions were carefully established based on corroborating evidence from several sources, including blood-and brain-based toxicology screens with confirmation by next-of-kin reports. Therefore, misclassification is unlikely. Comorbidities were also minimal, as cases and controls were drawn from decedents with no psychiatric or substance use disorder diagnoses, brain trauma, metastatic cancer, or communicable disease based on the determination of two independent board-certified psychiatrists upon review of abstracted medical records and next-of-kin reporting. We used the 2DF test, which achieves similar power to a standard 1DF test when no interaction is present and simultaneously improves power when interaction effects are present.^37^ Although we used conservative thresholds to identify the strongest interactions and the most robust signals, the full 2DF results are provided for researchers to explore additional signals, which are likely present. Finally, we tested colocalization across QTL data types with variants stemming from GSCAN’s seminal GWAS for cigarette smoking behaviors. To our knowledge, the present study represents the first large-scale formal testing of colocalization for GSCAN-identified loci with QTLs in human brain, and its results identified novel target genes and provided insight into the neurobiological function of smoking-associated heritable factors in relation to both DNAm and gene expression.

Overall, this multi-ancestry, multi-omics study of decedents with smoking status known and accounted for provides a unique resource for interrogating regions across the genome for their influence on gene regulation, cigarette smoking behaviors, and other complex conditions involving the NAc. Future studies may use these data to compare QTLs across other brain tissues to gain insights into tissue-specific regulation and to further investigate the neurobiology underlying other disease processes.

## Data Availability

All gene expression and DNA methylation data used in this study was previously generated and are available online through the Gene Expression Omnibus database accession GSE171936.

## Acknowledgements

This work was supported by the National Institute on Drug Abuse (NIDA) grants R01 DA042090 and R01 DA051913 to DBH and R21 DA051921 to HW. We gratefully thank the families who donated tissue to make this research possible. We also thank the Office of the Chief Medical Examiner of the State of Maryland, the Department of Pathology at Western Michigan University Homer Stryker MD School of Medicine, the University of North Dakota School of Medicine and Health Sciences, Department of Pathology, the County of Santa Clara Medical Examiner-Coroner Office, and the National Institute of Mental Health (NIMH) Intramural Research Program for their collaboration on tissue collection.

## References

1. GTEx Consortium. The GTEx Consortium atlas of genetic regulatory effects across human tissues. Science 369, 1318–1330 (2020).

2. Vosa, U. et al. Large-scale cis-and trans-eQTL analyses identify thousands of genetic loci and polygenic scores that regulate blood gene expression. Nat Genet 53, 1300–1310 (2021).

3. Perzel Mandell, K.A., et al. Genome-wide sequencing-based identification of methylation quantitative trait loci and their role in schizophrenia risk. Nat Commun 12, 5251 (2021).

4. Min, J.L. et al. Genomic and phenotypic insights from an atlas of genetic effects on DNA methylation. Nat Genet 53, 1311–1321 (2021).

5. Markunas, C.A., Johnson, E.O. & Hancock, D.B. Comprehensive evaluation of disease-and trait-specific enrichment for eight functional elements among GWAS-identified variants. Hum Genet 136, 911–919 (2017).

6. Gamazon, E.R. et al. Using an atlas of gene regulation across 44 human tissues to inform complex disease-and trait-associated variation. Nat Genet 50, 956–967 (2018).

7. Barbeira, A.N. et al. Exploiting the GTEx resources to decipher the mechanisms at GWAS loci. Genome Biol 22, 49 (2021).

8. Oliva, M. et al. The impact of sex on gene expression across human tissues. Science 369(2020).

9. Joehanes, R. et al. Epigenetic Signatures of Cigarette Smoking. Circ Cardiovasc Genet 9, 436–447 (2016).

10. Markunas, C.A. et al. Genome-wide DNA methylation differences in nucleus accumbens of smokers vs. nonsmokers. Neuropsychopharmacology 46, 554–560 (2021).

11. Semick, S.A. et al. Developmental effects of maternal smoking during pregnancy on the human frontal cortex transcriptome. Mol Psychiatry 25, 3267–3277 (2020).

12. Wang, D. et al. Comprehensive functional genomic resource and integrative model for the human brain. Science 362(2018).

13. Xiong, X. et al. Genetic drivers of m(6)A methylation in human brain, lung, heart and muscle. Nat Genet 53, 1156–1165 (2021).

14. Ng, B. et al. An xQTL map integrates the genetic architecture of the human brain’s transcriptome and epigenome. Nat Neurosci 20, 1418–1426 (2017).

15. Hancock, D.B. et al. A multiancestry study identifies novel genetic associations with CHRNA5 methylation in human brain and risk of nicotine dependence. Hum Mol Genet 24, 5940–54 (2015).

16. Wang, J.C. et al. Cis-regulatory variants affect CHRNA5 mRNA expression in populations of African and European ancestry. PLoS One 8, e80204 (2013).

17. Hancock, D.B. et al. Genome-wide association study across European and African American ancestries identifies a SNP in DNMT3B contributing to nicotine dependence. Mol Psychiatry 23, 1911–1919 (2018).

18. Liu, M. et al. Association studies of up to 1.2 million individuals yield new insights into the genetic etiology of tobacco and alcohol use. Nat Genet 51, 237–244 (2019).

19. Saunders, G.R.B. et al. Genetic diversity fuels gene discovery for tobacco and alcohol use. Nature 612, 720–724 (2022).

20. Koob, G.F. & Volkow, N.D. Neurobiology of addiction: a neurocircuitry analysis. Lancet Psychiatry 3, 760–773 (2016).

21. Grace, A.A., Floresco, S.B., Goto, Y. & Lodge, D.J. Regulation of firing of dopaminergic neurons and control of goal-directed behaviors. Trends Neurosci 30, 220–7 (2007).

22. BrainSeq, A.H.B.G.C. BrainSeq: Neurogenomics to Drive Novel Target Discovery for Neuropsychiatric Disorders. Neuron 88, 1078–1083 (2015).

23. Avila-Tang, E. et al. Assessing secondhand smoke using biological markers. Tob Control 22, 164–71 (2013).

24. Anderson, C.A. et al. Data quality control in genetic case-control association studies. Nat Protoc 5, 1564–73 (2010).

25. Delaneau, O., Howie, B., Cox, A.J., Zagury, J.F. & Marchini, J. Haplotype estimation using sequencing reads. Am J Hum Genet 93, 687–96 (2013).

26. Marchini, J., Howie, B., Myers, S., McVean, G. & Donnelly, P. A new multipoint method for genome-wide association studies by imputation of genotypes. Nat Genet 39, 906–13 (2007).

27. Numata, S. et al. DNA methylation signatures in development and aging of the human prefrontal cortex. Am J Hum Genet 90, 260–72 (2012).

28. Jaffe, A.E. et al. Mapping DNA methylation across development, genotype and schizophrenia in the human frontal cortex. Nat Neurosci 19, 40–7 (2016).

29. Aryee, M.J. et al. Minfi: a flexible and comprehensive Bioconductor package for the analysis of Infinium DNA methylation microarrays. Bioinformatics 30, 1363–9 (2014).

30. Jaffe, A.E. et al. Developmental regulation of human cortex transcription and its clinical relevance at single base resolution. Nat Neurosci 18, 154–161 (2015).

31. Houseman, E.A. et al. DNA methylation arrays as surrogate measures of cell mixture distribution. BMC Bioinformatics 13, 86 (2012).

32. Jaffe, A.E. et al. Developmental and genetic regulation of the human cortex transcriptome illuminate schizophrenia pathogenesis. Nat Neurosci 21, 1117–1125 (2018).

33. Bolger, A.M., Lohse, M. & Usadel, B. Trimmomatic: a flexible trimmer for Illumina sequence data. Bioinformatics 30, 2114–20 (2014).

34. Patro, R., Duggal, G., Love, M.I., Irizarry, R.A. & Kingsford, C. Salmon provides fast and bias-aware quantification of transcript expression. Nat Methods 14, 417–419 (2017).

35. Srivastava, A. et al. Alignment and mapping methodology influence transcript abundance estimation. Genome Biol 21, 239 (2020).

36. Soneson, C., Love, M.I. & Robinson, M.D. Differential analyses for RNA-seq: transcript-level estimates improve gene-level inferences. F1000Res 4, 1521 (2015).

37. Aschard, H., Hancock, D.B., London, S.J. & Kraft, P. Genome-wide meta-analysis of joint tests for genetic and gene-environment interaction effects. Hum Hered 70, 292–300 (2010).

38. Love, M.I., Huber, W. & Anders, S. Moderated estimation of fold change and dispersion for RNA-seq data with DESeq2. Genome Biol 15, 550 (2014).

39. Stegle, O., Parts, L., Piipari, M., Winn, J. & Durbin, R. Using probabilistic estimation of expression residuals (PEER) to obtain increased power and interpretability of gene expression analyses. Nat Protoc 7, 500–7 (2012).

40. Huang, Q.Q., Ritchie, S.C., Brozynska, M. & Inouye, M. Power, false discovery rate and Winner’s Curse in eQTL studies. Nucleic Acids Res 46, e133 (2018).

41. Davis, J.R. et al. An Efficient Multiple-Testing Adjustment for eQTL Studies that Accounts for Linkage Disequilibrium between Variants. Am J Hum Genet 98, 216–24 (2016).

42. Subramanian, A. et al. Gene set enrichment analysis: a knowledge-based approach for interpreting genome-wide expression profiles. Proc Natl Acad Sci U S A 102, 15545–50 (2005).

43. Pers, T.H., Timshel, P. & Hirschhorn, J.N. SNPsnap: a Web-based tool for identification and annotation of matched SNPs. Bioinformatics 31, 418–20 (2015).

44. Foley, C.N. et al. A fast and efficient colocalization algorithm for identifying shared genetic risk factors across multiple traits. Nat Commun 12, 764 (2021).

45. Chen, F. et al. Multi-ancestry transcriptome-wide association analyses yield insights into tobacco use biology and drug repurposing. Nat Genet 55, 291–300 (2023).

46. Cornelius, M.E., Wang, T.W., Jamal, A., Loretan, C.G. & Neff, L.J. Tobacco Product Use Among Adults -United States, 2019. *MMWR Morb Mortal Wkly Rep* **69**, 1736-1742 (2020).

47. Organization;, W.H. WHO report on the global tobacco epidemic. (2017).

48. Koob, G.F. & Volkow, N.D. Neurocircuitry of addiction. *Neuropsychopharmacology* **35**, 217–38 (2010).

49. Oliva, M. et al. DNA methylation QTL mapping across diverse human tissues provides molecular links between genetic variation and complex traits. Nat Genet (2022).

50. McKay, B.E., Placzek, A.N. & Dani, J.A. Regulation of synaptic transmission and plasticity by neuronal nicotinic acetylcholine receptors. Biochem Pharmacol 74, 1120–33 (2007).

51. Siedlinski, M. et al. Genome-wide association study of smoking behaviours in patients with COPD. Thorax 66, 894–902 (2011).

52. Yang, J. et al. Chronic nicotine differentially affects murine transcriptome profiling in isolated cortical interneurons and pyramidal neurons. BMC Genomics 18, 194 (2017).

53. Gill, K.J. & Boyle, A.E. Genetic basis for the psychostimulant effects of nicotine: a quantitative trait locus analysis in AcB/BcA recombinant congenic mice. Genes Brain Behav 4, 401–11 (2005).

54. Baker, E.J., Jay, J.J., Bubier, J.A., Langston, M.A. & Chesler, E.J. GeneWeaver: a web-based system for integrative functional genomics. Nucleic Acids Res 40, D1067–76 (2012).

55. Gelernter, J. et al. Genome-wide association study of cocaine dependence and related traits: FAM53B identified as a risk gene. Mol Psychiatry 19, 717–23 (2014).

56. Lee, M.K., Hong, Y., Kim, S.Y., London, S.J. & Kim, W.J. DNA methylation and smoking in Korean adults: epigenome-wide association study. Clin Epigenetics 8, 103 (2016).

57. Nannini, D.R. et al. Genome-wide DNA methylation association study of recent and cumulative marijuana use in middle aged adults. Mol Psychiatry (2023).

58. Fuemmeler, B.F. et al. DNA Methylation in Babies Born to Nonsmoking Mothers Exposed to Secondhand Smoke during Pregnancy: An Epigenome-Wide Association Study. Environ Health Perspect 129, 57010 (2021).

59. Shorey-Kendrick, L.E. et al. Impact of vitamin C supplementation on placental DNA methylation changes related to maternal smoking: association with gene expression and respiratory outcomes. Clin Epigenetics 13, 177 (2021).

60. Lin, H. et al. Prefrontal cortex eQTLs/mQTLs enriched in genetic variants associated with alcohol use disorder and other diseases. Epigenomics 12, 789–800 (2020).

61. Pierce, B.L. et al. Co-occurring expression and methylation QTLs allow detection of common causal variants and shared biological mechanisms. Nat Commun 9, 804 (2018).

62. Hadjiargyrou, M. Mustn1: A Developmentally Regulated Pan-Musculoskeletal Cell Marker and Regulatory Gene. Int J Mol Sci 19(2018).

63. Human Protein Atlas. Vol. 2023.

64. Uhlen, M. et al. Proteomics. Tissue-based map of the human proteome. Science 347, 1260419 (2015).

65. Ohi, K. et al. Schizophrenia risk variants in ITIH4 and CALN1 regulate gene expression in the dorsolateral prefrontal cortex. Psychiatr Genet 26, 142–3 (2016).

66. Bandow, J.E. et al. Improved image analysis workflow for 2-D gels enables large-scale 2-D gel-based proteomics studies--COPD biomarker discovery study. Proteomics 8, 3030–41 (2008).

67. Lee, K.Y. et al. Inter-alpha-trypsin inhibitor heavy chain 4: a novel biomarker for environmental exposure to particulate air pollution in patients with chronic obstructive pulmonary disease. Int J Chron Obstruct Pulmon Dis 10, 831–41 (2015).

68. Delezie, J. & Handschin, C. Endocrine Crosstalk Between Skeletal Muscle and the Brain. Front Neurol 9, 698 (2018).

69. Isaac, A.R., Lima-Filho, R.A.S. & Lourenco, M.V. How does the skeletal muscle communicate with the brain in health and disease? Neuropharmacology 197, 108744 (2021).

