## Supplemental Materials for "Smoking-informed methylation and expression QTLs in human brain and colocalization with smoking-associated genetic loci"

### Supplementary Materials

### Supplemental Methods

#### Human Postmortem NAc Samples

Postmortem human NAc tissues were obtained from the Lieber Institute for Brain Development (LIBD) brain collection at autopsy, as previously described.[1,2] Decedents with DSM-5 psychiatric or substance use disorders other than nicotine were excluded in addition to those with brain trauma, metastatic brain cancer, neurotic pathology, neurodegenerative diseases, HIV/AIDS, hepatitis, or other communicable diseases. Information regarding demographics, substance use history, and current smoking status were collected from a next-of-kin 36-item telephone-administered questionnaire (LIBD Autopsy Questionnaire). Donor information was also vetted using medical examiner documents and medical records compiled into a narrative summary, which was reviewed by two psychiatrists. Cotinine biomarker measures from brain and/or blood samples were measured using a standard toxicology screen (National Medical Services Labs, Inc., Willow Grove, Pennsylvania). As used in Markunas et al., smoking cases were defined by cotinine levels above 12 ng/mL in blood and 12 ng/g in brain, a threshold that differentiates between active and passive smoking,[3] and a next-of-kin report of current smoking.[1] Controls (nonsmokers) were defined by cotinine levels below 12 ng/mL (blood) or 12 ng/g (brain) and a next-of-kin report of no current smoking.

#### Genotype Data

The genotype data in this study were obtained from a superset of samples genotyped and imputed as part of the full LIBD cohort, using previously described procedures.[2] Briefly, samples were genotyped on Illumina microarrays (HumanHap650 [27%], Human1M [35%], HumanOmni2.5 [19%], or HumanOmni5-Quad [19%] BeadChips among the samples included in the present study), and a standard quality control (QC) protocol[4] was followed to remove low-quality (Hardy-Weinberg equilibrium p-value $<1\times10^{-6}$) and low-frequency (minor allele frequency [MAF] <0.005) variants for each separate genotyping array. Haplotypes were phased using SHAPEIT,[5] and then genome-wide imputation was conducted, separately by genotyping array, using IMPUTE2[6] with reference to 1000 Genomes phase 3. Imputed genotype dosages were converted to hard-call genotypes for variants with imputation posterior probabilities >0.9. For each genotyping array, the hard-call genotypes for the corresponding sample subset were iteratively merged with the Human1M sample subset using qctool (https://www.well.ox.ac.uk/~gav/qctool/). The aforementioned standard QC filtering was repeated on the post-imputation, hard-call genotype data in the full LIBD cohort.[2] In the subset included in the present study, over 11 million autosomal variants were carried forward for analyses.

#### DNAm and RNA-seq Data Generation and Processing

DNA and RNA were extracted from NAc samples of 239 eligible decedents, as described previously.[7,8] DNAm was measured using an Illumina Human MethylationEPIC BeadChip. As described before,[1] DNAm data processing was conducted using the R package *minfi*,[9] which included stratified quantile normalization, correcting technical artifacts using principal components (PCs) of the negative control probe intensities, and controlling for tissue sample heterogeneity by estimating neuronal cell-type proportions[10] using the Houseman method.[11] DNAm β-values were calculated and used in the meQTL analyses, representing the percentage of DNAm at each CpG (ratio of methylated intensities relative to the total intensity).

Following RNA extraction protocols previously described, samples were sequenced using paired-end 100 bp reads on an Illumina HiSeq3000 at LIBD.[1,12] For each RNA-seq sample, reads were trimmed and filtered using *Trimmomatic v0.39.*[13] Adapter sequences were trimmed if detected, and low-quality reads or read ends were removed using the following settings: *ILLUMINACLIP seedMismatches=2*, *ILLUMINACLIP palindromeClipThreshold=30*, *ILLUMINACLIP simpleClipThreshold=10*, *ILLUMINACLIP minAdapterLength=4*, *ILLUMINACLIP keepBothReads=true*, *LEADING quality=3*, *TRAILING quality=3*, *SLIDINGWINDOW windowSize=4*, *SLIDINGWINDOW requiredQuality=10*, *MINLEN length=75*, *AVGQUAL quality=20*. For transcript quantification, read pairs with both reads passing quality filtering were pseudo-mapped using *Salmon v1.1.0*[14] in selective alignment mode[15] using the GENCODE v34 (ENSEMBL release 100) comprehensive gene annotation as the transcriptome index and the full GRCh38 primary genome assembly as a selective alignment decoy sequence. The use of selective alignment enhances the accuracy of transcript quantification by accounting for the possibility of intronic and intergenic DNA sequence artifacts in the sequenced libraries. The *--gcBias* and *--seqBias* options were enabled by default to account for other potential transcript quantification biases.[16] *Salmon* transcript quantifications were aggregated to the gene-level counts using *tximport v1.12.3*,[17] resulting in 60,240 GENCODE genes. To generate QC metrics and mapping statistics for read alignment, reads were aligned to the GRCh38 primary genome assembly using *HISAT2 v2.1.0*.[18] Quality metrics for raw and post-*Trimmomatic* reads were generated using *FASTQC v0.11.8* (https://www.bioinformatics.babraham.ac.uk/projects/fastqc/). All metrics and statistics were aggregated using *MultiQC v1.7*.[19]

To identify potential sample swaps, genotype correlations derived from 1000 Genomes Phase 3 imputed genotype and RNA-seq data were calculated. Genotypes were called from RNA-seq data using the mpileup utility from SAMTools.[20] Problematic samples were identified by calculating pairwise genotype correlations (Pearson correlation coefficient ρ) between RNA-seq and genotype datasets. Samples were either excluded or mismatches resolved if $\rho>0.8$ for non-matching sample IDs or $\rho<0.7$ for matching sample IDs. Similar QC was conducted using the DNAm data [1].

Samples were excluded based on the following criteria: effective sequencing depth<10 million read pairs; transcriptome mapping rate<0.3; intergenic mapping rate>0.3 when transcriptome mapping rate<0.5; transcript diversity (i.e., Shannon index) outlier when transcriptome mapping rate<0.5; mitochondrial mapping rate>0.2; ribosomal RNA mapping rate>0.01; RIN score<6; missing self-reported sex data; chromosome Y gene expression-based sex discordance with reported sex; missing genotype data; and problematic genotype correlations. Sample-level QC removed 36 samples, resulting in a post-QC RNA-seq sample size of 203. Lowly expressed genes were then removed using the exclusion criteria of ≥90% of samples with ≤10 gene counts or ≤1 transcripts per million value. Because the genetic variants for eQTL mapping were annotated to the GRCh37 human genome reference, gene annotations were represented as GRCh37 coordinates, resulting in a final set of 16,274 genes considered for eQTL mapping.

Of the available 239 decedents, 201 RNA-seq samples and 220 DNAm samples with genotype and smoking data remained following QC, including 198 samples in both datasets (intersection) and 223 samples with either RNA-seq or DNAm data (union).

#### Methylation and Expression Quantitative Trait Loci (meQTL/eQTL) Mapping

We performed *cis*-DNAm quantitative trait loci (meQTL) mapping using imputed genetic variants and DNAm intensity β-values of probes proximal (within 500 kb up or downstream) to these variants. Four different meQTL mapping models were fit: (1) A "baseline" model to test for association between genetic variants and DNAm β-values across both smoking cases and controls; (2) a smoking cases–only model similar to the baseline model; (3) a smoking controls–only model similar to the baseline model; and (4) an interaction model to test for associations of a genetic variant-by-smoking status interaction with DNAm β-values. The smoking cases–only and controls-only models are needed to generate summary statistics used to conduct stratified two degrees-of-freedom (2DF) tests. The stratified 2DF test jointly tests for genetic variant main effects and genetic variant-by-smoking status interaction effects.[21] For each genetic variant and DNAm probe pair, the stratified 2DF test used $t$-statistics for the genetic variant effect ($\gamma$ in Equation 1 below) for the smoking cases–only and controls-only models. The $t$-statistics were squared then added together to form a $\chi^2$ statistic from which a two-sided p-value was derived.

For the baseline, smoking cases–only, and smoking controls–only meQTL mapping, we fit the following linear model for each DNAm probe using Matrix eQTL v2.2[22]:

$$Y = \beta_0 + \gamma * g + \sum_1^k \beta_i * x_i + \varepsilon \qquad (1)$$

Here, $Y$ is the rank-inverse normal transformed (RINT) DNAm β-value for a given CpG probe, $\beta_0$ is the intercept term, $g$ is the imputed hard-call genotype for a given genetic variant, $x_i$ are technical and biological variables to account for potential confounding, and $\varepsilon$ is the error term. Technical and biological variables included age at death, sex, estimated non-neuronal cell-type proportion, PC1 for DNAm array negative control probes, PC1 for imputed genotypes, and PC2 for imputed genotypes. These variables were selected from a larger list of variables to obtain a parsimonious model that reduced model complexity while accounting for potential confounding. Pairwise correlations between all variables were evaluated to determine and exclude variables that represented redundant information (e.g., genotype PC1 was included in lieu of self-reported race due to strong correlation). Additionally, variables were assessed for significant association with smoking status by fitting logistic regression models that included smoking status as the outcome variable and potential meQTL model covariates as predictor variables. No significant associations with smoking status were identified based on two-sided t-tests for the regression coefficients. Likewise, the interaction model was fit using Matrix eQTL:

$$Y = \beta_0 + \gamma * g + \sum_1^{k-1} \beta_i * x_i + \delta * g * x_k + \varepsilon \qquad (2)$$

$Y$, $g$, and $x_1, x_2, \ldots, x_{k-1}$ are the same as in Equation 1, and $x_k$ is the smoking status variable. The $\delta$ term is the estimated genetic variant-by-smoking interaction effect.

A similar framework to meQTL mapping was applied for *cis*-expression quantitative trait loci (eQTL) mapping. Baseline, smoking cases–only, smoking controls–only, and interaction eQTL models were fit for each imputed genetic variant and NAc expression levels of genes proximal (within 500 kb of gene body) to these variants. The general structure of Equation 1 was used for the baseline and smoking status stratified eQTL mapping models, and the general structure of Equation 2 was used for the interaction model. Here, $Y$ is the count values for a given gene after median-of-ratios normalization[23] and RINT. The biological and technical variables denoted by $x_i$ were age at death, sex, exon mapping rate, ribosomal RNA mapping rate, PC1 for imputed genotypes, PC2 for imputed genotypes, and four latent variables estimated by

PEER v1.3[24] to account for additional unmeasured sources of confounding. PEER factors (i.e., the latent variables) were estimated using counts after median-of-ratios normalization and the *DESeq2 v1.26.0* variance stabilizing transformation, *vst*. Covariates input to PEER during factor estimation include smoking status, age at death, sex, exon mapping rate, ribosomal RNA mapping rate, PC1 for imputed genotypes, and PC2 for imputed genotypes. Initially, the top five PEER factors, based on the posterior variance of factor weights, were selected as explanatory variables for the eQTL models. However, the top PEER factor was excluded due to strong correlation with the exon mapping rate (Spearman correlation coefficient=0.99). Model selection for the eQTL mapping was conducted similarly to as described for the meQTL mapping model selection.

For the purposes of QTL mapping, we defined "proximal" as a maximum distance of 500 kb from genetic variants to DNAm probes and genes. This window size was chosen by applying Equation 1 for meQTL mapping on multiple chromosomes using a larger *cis*-window size of 2 Mb (1 Mb up and downstream of the DNAm probe). We observed a systematic decrease in genetic variant–DNAm probe association $-\log_{10}$ p-values with increasing variant distance from a DNAm probe. Using the approximate inflection point for $-\log_{10}$ p-values as a function of variant distance from a DNAm probe, we selected a total *cis*-window size of 1 Mb (500 kb upstream to 500 kb downstream).

To account for multiple hypothesis testing in the meQTL/eQTL baseline and interaction models and stratified 2DF tests, p-values were corrected using a two-stage approach to mitigate inflation associated with single-stage approaches.[25] This hierarchical approach first accounts for association tests across variants for a given DNAm probe/gene, then accounts for tests across all probes/genes. In the first stage, all nominal p-values for a given probe/gene were adjusted using EigenMT[26] with parameters *--var_thresh 0.99*, *--window 200*, and *--cis_dist 500000*. In the second stage, EigenMT adjusted p-values were further corrected by the number of probes/genes tested. Any initial two-stage adjusted p-value smaller than a Bonferroni corrected p-value was assigned the latter p-value as the final two-stage, adjusted p-value. This ensures that the two-stage adjustment stringency does not exceed the family-wise error rate control of Bonferroni correction. A two-stage adjusted p-value cutoff of 0.05 was used to identify statistically significant QTLs.

##### Gene Set Enrichment Analysis (GSEA) for eGenes and meQTL CpGs

We used the GSEAPreranked tool from GSEA v4.3.2[27] to test for enrichment of smoking-associated eGenes and meQTL CpGs in MSigDB v2023.1.Hs gene set collections *C2 canonical pathways* and *C5 Gene Ontology gene sets*. Only gene sets with 15–500 genes were included. CpGs were assigned to genes using the Infinium MethylationEPIC v1.0 B5 Manifest, resulting in 11,772 CpGs from the interaction meQTL mapping model with at least one assigned gene. Each gene was ranked using the largest $-\log_{10}$-transformed smoking interaction p-value for a given gene from QTL mapping. Weighted Kolmogorov-Smirnov–like statistics were computed for the enrichment scores and p-values were determined using 1,000 gene set permutations. Significant pathways were selected based on an FDR threshold of 0.10.

##### QTL enrichment testing for GSCAN genetic variants

We conducted variant-based enrichment testing to assess whether genome-wide significant GSCAN loci were enriched for meQTLs or eQTLs. We obtained GSCAN summary statistics from University of Minnesota's Data Repository for U of M (https://doi.org/10.13020/3b1n-ff32), focusing on GSCAN's GWAS results from 2019 to capture genetic loci with variants that commonly occur and have the largest effect sizes on smoking: N up to 1.2 million individuals, depending on the smoking trait analyzed.[28] For meQTL enrichment analysis, we compared the p-value distributions from stratified 2DF tests (i.e., stratified 2DF meQTL mapping) between GSCAN variants and a set of randomly matched variants. The GSCAN variant set included linkage disequilibrium (LD)-pruned, genome-wide significant variants reported by GSCAN, across all four smoking traits, that were also available in our meQTL mapping (361 variants). LD pruning was implemented using PLINK v2.0[29] with the parameterss *--indep-pairwise 1500 150 0.2*. The matched variant set included 3,600 LD-pruned variants selected using SNPsnap.[30] SNPsnap constructed a set of randomly drawn autosomal variants from a larger set of variants matched to GSCAN-reported variants based on (1) MAFs, (2) number of variants in LD, (3) distance to nearest gene, and (4) gene density. For variants with multiple stratified 2DF p-values (i.e., proximal to multiple CpG sites), only the smallest p-value was retained. The meQTL stratified 2DF p-value distributions for the GSCAN and matched variant sets were tested for equality using a two-sided Kolmogorov-Smirnov test. The eQTL enrichment analysis followed the same procedure as the meQTL enrichment analysis. The GSCAN variant set included 305 variants because the overlap with variants from eQTL mapping differed from meQTL mapping. The SNPsnap-constructed matched variant set included 3,050 variants.

##### Colocalization between GSCAN GWAS and meQTL/eQTL mappings

We tested whether meQTL or eQTL signals from the baseline model QTL mapping colocalized with GSCAN loci for smoking initiation, age at initiation, cessation, and cigarettes per day using the *coloc v5.1.0* R package. For simplicity, we present the details of this analysis in relation to a single GSCAN trait and meQTLs. An equivalent framework was applied for GSCAN–eQTL colocalization testing. For GSCAN–meQTL colocalization analysis, we defined genomic regions to test based on a multi-step selection process. For a given GSCAN locus, DNAm probes were obtained that had at least one variant–CpG association test for a variant within the locus. For each of these DNAm probes, all variants that were tested for association with the probe in the baseline model meQTL mapping and were also tested in the GSCAN GWAS comprised the colocalization test region (Figure S1). Summary statistics from GSCAN and the baseline model meQTL mapping for these variants were provided to the *coloc.abf* function to perform colocalization testing. Parameter setting *type="quant"* was specified for age of initiation, cigarettes per day, and DNAm probe intensity values, and *type="cc"* was specified for smoking initiation and smoking cessation. From the GSCAN summary statistics, *coloc.abf* was supplied the regression coefficients (i.e., betas), coefficient standard errors (SEs), sample sizes, minor allele frequencies, and proportions of cases for the case-control (*type="cc")* traits. From the baseline model meQTL mapping summary statistics, we supplied the regression coefficients (i.e., betas), coefficient SEs, sample size (220 for meQTL mapping and 201 for eQTL mapping), and standard deviations of the RINT DNAm probe intensity values. A colocalization test region was considered as having a colocalized signal between the GSCAN GWAS and meQTL mapping if the *coloc* posterior probability of hypothesis 4 (both traits are associated and share a single causal variant) exceeded 0.8, and the meQTL had a genome-wide significant two-stage adjusted p-value in the baseline meQTL analysis.

For GSCAN loci that showed colocalization with both an meQTL and an eQTL, HyPrColoc (https://github.com/jrs95/hyprcoloc/; commit ID f279ceb) was applied to assess whether these colocalizations resulted from the same region of the locus.[31] Baseline model QTL mapping summary statistics for all eGenes and CpGs that independently colocalized with a given GSCAN locus were used to obtain all variant–eGene and variant–CpG regression coefficients (i.e., genetic variant effects on gene expression/DNA methylation) and coefficient SEs. Likewise, GSCAN summary statistics were used to obtain coefficients and SEs for genetic variant associations with the given GSCAN trait at the given genomic locus. Each HyPrColoc test only included one GSCAN trait, eGene, and CpG, so if multiple genes or CpGs independently colocalized with the GSCAN trait, all combinations of gene–CpG pairs were combined with the GSCAN trait for a HyPrColoc test. For each HyPrColoc test, all genetic variants that had summary statistics available across the GSCAN trait, eGene, and CpG were provided as input to HyPrColoc. Significant colocalization was defined as a GSCAN trait–eGene–CpG triplet having posterior probability >0.8.

#### Annotation of meQTL and eQTL

For the supplemental tables that report meQTL and eQTL summary statistics, variants were annotated using SNPnexus,[32] a web-based variant annotation tool. Annotations and variant position are based on the UCSC genome browser database (genome build hg19). Intragenic variants were annotated to the genes in which they reside, and intergenic variants were annotated to their nearest genes (multiple genes are separated by the delimiter "|"). Table cell values of "-" for variant-to-gene mappings indicate that the variant is located >1.5 kilobases (kb) upstream of the nearest gene transcription start site (TSS) or >1.5 kb downstream of the polyadenylation signal.

CpGs were annotated based on UCSC hg19 RefGene data in the Infinium MethylationEPIC v1.0 B5 Manifest File (https://support.illumina.com/downloads/infinium-methylationepic-v1-0-product-files.html). Table cell values of "-" for CpG-to-gene mappings denotes CpGs for which the nearest gene is located >1.5 kb upstream of the nearest gene transcription start site (TSS) or >1.5 kb downstream of the polyadenylation signal. Category definitions for a CpG's location in relation to its nearest gene(s) are defined below:

- 5' UTR: within the 5' untranslated region (UTR) between the TSS and the ATG start site.
- Body: between the ATG and stop codon irrespective of the presence of introns, exons, TSS, or promoters.
- TSS200: 0–200 bases upstream of the TSS.
- TSS1500: 200–1,500 bases upstream of the TSS.
- 3' UTR: Between the stop codon and polyadenylation signal.

The eQTL-associated genes (eGenes) were annotated using the GENCODE v34 (ENSEMBL release 100) primary assembly comprehensive gene annotation, genome build hg19. Summary statistics and annotations for Genotype-Tissue Expression (GTEx) project eQTL were obtained from publicly available nucleus accumbens and basal ganglia GTEx v8 eQTL maps (https://www.gtexportal.org/home/datasets).

### Supplemental Figures

**Supplemental Figure S1. Selection of GWAS-identified regions for colocalization testing using *coloc*.**

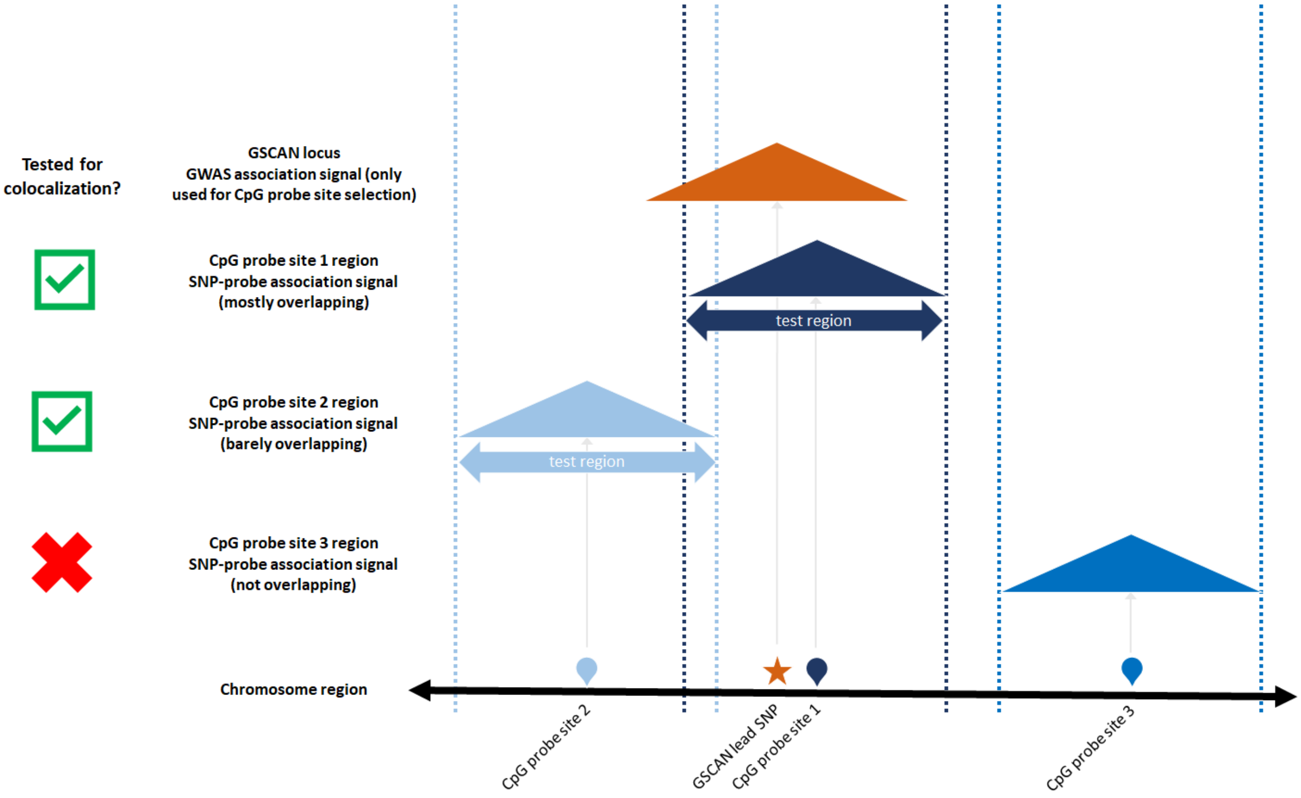


This diagram illustrates two examples of CpGs that meet inclusion criteria for colocalization analysis and one example of a CpG that would not be included. For a given GSCAN-reported significant GWAS locus for one of the smoking phenotypes (smoking initiation, age of initiation, cigarettes per day, or cessation), colocalization analysis was conducted with a CpG if any variants tested for association with the CpG (i.e., meQTL mapping) overlapped with the GSCAN locus. The colocalization test region included the intersection set of variants in the *cis*-meQTL mapping for the CpG that were also tested in the GSCAN GWAS. As illustrated, the size of the test region that overlapped with the GSCAN locus was CpG-dependent. A similar procedure was applied for colocalization testing of GSCAN loci with genes considered in eQTL mapping.

**Supplemental Figure S2. Top 5 meQTLs with evidence of an interaction with smoking.**

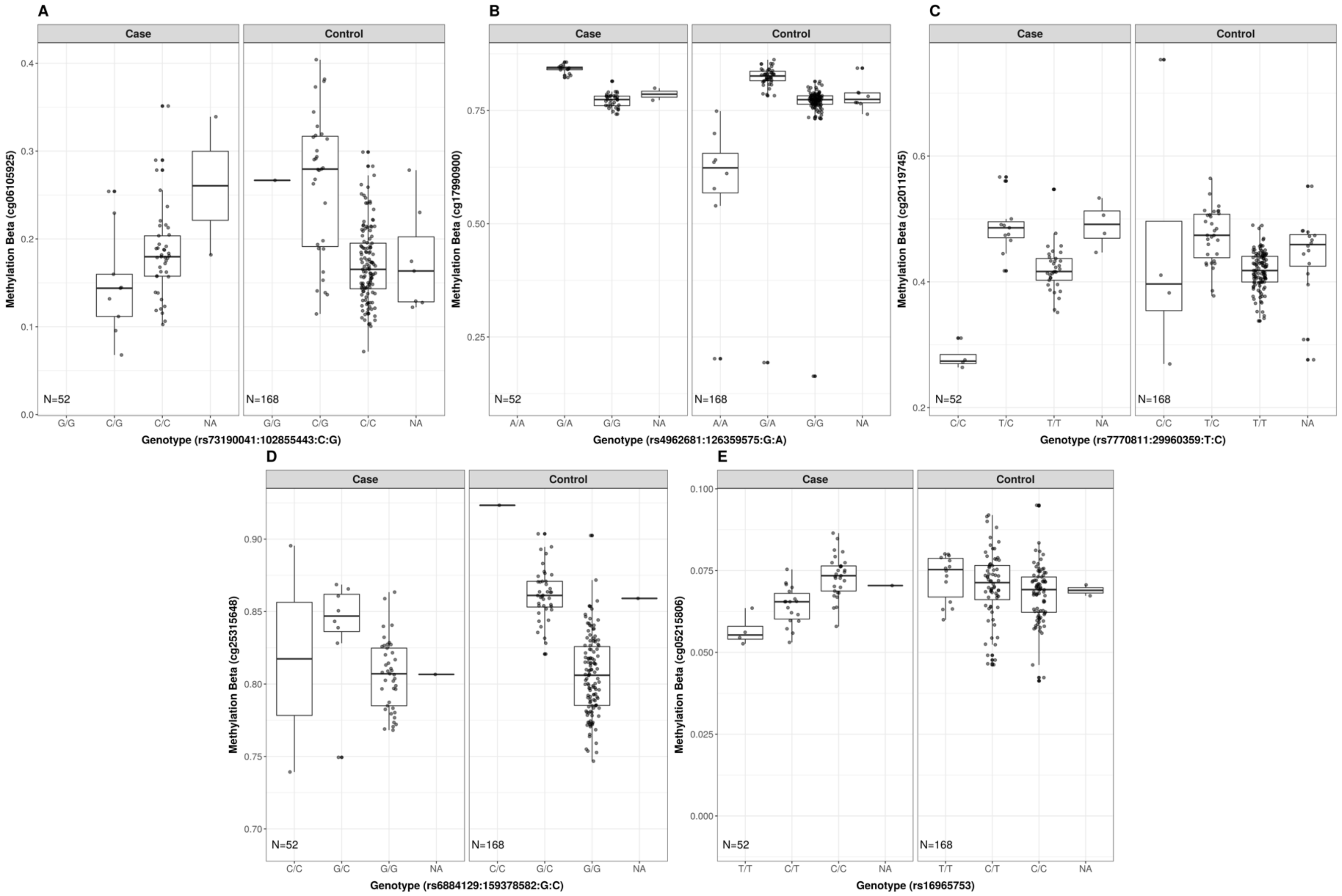


Boxplots showing the relationship between genotype (x-axes) and DNAm expression values (y-axes) straitified by smoking status for the five meQTLs presented in Table 2 (meQTLs with the strongest genotype smoking status interactions by p-value ranking that pass MAF and missingness filters in both EA and AA). Each plot point represents a sample, and missing genotypes are denoted by "NA" values. Matrix eQTL imputes missing genotypes to the mean.

### Supplemental Table Legends

**Supplemental Table S1. Genome-wide significant NAc *cis*-meQTL lead variants**. Each row denotes the lead variant from a significant *cis*-meQTL (adjusted p-value$_{2DF}$ <0.05). Lead variants are defined as the genetic variant within an meQTL with the lowest stratified 2DF test p-value. In total, 784,843 CpGs were included in the meQTL mapping with 1,748,985,510 variant–CpG association tests conducted across these CpGs. Variants and CpGs corresponding to the significant meQTLs were annotated as described in the Supplemental Methods. The *cis*-meQTL summary statistics are presented for main (i.e., genetic variant) effects (baseline, smoking cases–only, and smoking controls–only meQTL models), interaction (i.e., variant-by-smoking status) effects, and joint 2DF tests. Details of each model are described in the Methods and Supplemental Methods. EA- and AA-specific variant metrics include ancestry-specific allele frequencies and missingness. For the combined MAF and missingness filter (column Q), values of "1" indicate a variant satisfying the conditions. Variants that did not meet these conditions were assigned a "0" value. Abbreviations: AA = African ancestry; bp = base pairs; EA = European ancestry; MAF = minor allele frequency; meQTL = methylation quantitative trait locus; n/a = not available; NAc = nucleus accumbens; SE = standard error; 2DF = 2 degrees-of-freedom.

**Supplemental Table S2**. **Genome-wide significant NAc *cis*-eQTLs**. Each row denotes a significant variant–gene association from *cis*-eQTL mapping (adjusted p-value$_{2DF}$ <0.05). Variants and genes corresponding to the eQTLs were annotated as described in the Supplemental Methods. The *cis*-eQTL summary statistics are presented for main (i.e., genetic variant) effects (baseline, smoking cases–only, and smoking controls–only eQTL models), interaction (i.e., variant-by-smoking status) effects, and joint 2DF tests. Details of each model are described in the Methods and Supplemental Methods. EA- and AA-specific variant metrics include ancestry-specific allele frequencies and missingness. For the combined MAF and missingness filter (column T), values of "1" indicate a variant satisfying the conditions. Variants that did not meet these conditions were assigned a "0" value. Abbreviations: AA = African ancestry; bp = base pairs; EA = European ancestry; eQTL = expression quantitative trait locus; MAF = minor allele frequency; n/a = not available; NAc = nucleus accumbens; SE = standard error; TSS = transcription start site; 2DF = 2 degrees-of-freedom.

**Supplemental Table S3**. **Genome-wide significant NAc *cis*-eQTL lead variants.** Each row denotes the lead variant from a significant *cis*-eQTL (adjusted p-value$_{2DF}$ <0.05). Lead variants are defined as the genetic variant within an eQTL with the lowest stratified 2DF test p-value. Columns are equivalent to Supplemental Table S2. Abbreviations: AA = African ancestry; bp = base pairs; EA = European ancestry; eQTL = expression quantitative trait locus; MAF = minor allele frequency; n/a = not available; NAc = nucleus accumbens; SE = standard error; TSS = transcription start site; 2DF = 2 degrees-of-freedom.

**Supplemental Table S4**. **GSEA Pathway Analysis.** Each row denotes a significant (defined by FDR q-value <0.1) gene set term from the Molecular Signatures Database (MSigDB) Canonical Pathways and Gene Ontology collections identified using the GSEA Preranked tool, ranking the meQTLs/eQTLs by smoking interaction evidence (-$\log_{10}$-transformed smoking interaction p-value). The *Rank at max* column denotes the position in the ranked GSEA Preranked input gene list at which the maximum enrichment score occurred. The *Leading edge* column provides statistics for the leading-edge subset, the subset of gene set genes that contribute most to the enrichment score. The *tags* statistic denotes the percentage of GSEA Preranked input genes contributing to the enrichment score. The *list* statistic indicates where in the ranked input gene list the enrichment score is attained. The *signal* statistic combines the tag and list statistics to indicate the density of gene set genes towards the beginning of the ranked input gene list.

**Supplemental Table S5. GSCAN meQTL Lookup *cis*-meQTL evidence among GSCAN GWAS-identified variants.** Each row corresponds to a significant GSCAN smoking phenotype GWAS variant that also has a significant variant–CpG association for an meQTL in our study. Significant variant–CpG associations are defined using a Bonferroni correction of stratified 2DF test p-values based on the set of significant GSCAN variants ($p\text{-value}_{2DF}<1.39\times10^{-4}$ [0.05/361 GSCAN significant variants that overlap with our *cis*-meQTL map]). Details on annotation of variants and CpGs are described in the Supplemental Methods. EA- and AA-specific variant metrics include ancestry-specific allele frequencies and missingness. The *cis*-meQTL summary statistics are presented for main (i.e., genetic variant) effects (baseline, smoking cases–only, and smoking controls–only meQTL models), interaction (i.e., variant-by-smoking status) effects, and joint 2DF tests. Details of each model are described in the Methods and Supplemental Methods. Abbreviations: AA = African ancestry; aai = age of initiation; cpd = cigarettes per day; EA = European ancestry; LIBD = Lieber Institute for Brain Development; meQTL = methylation quantitative trait locus; NAc = nucleus accumbens; sc = smoking cessation; si = smoking initiation; 2DF = 2 degrees-of-freedom.

**Supplemental Table S6**. **GSCAN–eQTL Lookup – *cis*-eQTL evidence among GSCAN GWAS-identified variants**. Each row corresponds to a significant GSCAN smoking phenotype GWAS variant that also has a significant variant–gene association for an eQTL in our study. Significant variant–gene associations are defined using a Bonferroni correction of stratified 2DF test p-values based on the set of significant GSCAN variants ($p\text{-value}_{2DF}<1.66\times10^{-4}$ [0.05/305 GSCAN significant variants that overlap with our *cis*-eQTL map]). Details on annotation of variants are described in the Supplemental Methods. EA- and AA-specific variant metrics include ancestry-specific allele frequencies and missingness. The *cis*-eQTL summary statistics are presented for main (i.e., genetic variant) effects (baseline, smoking cases–only, and smoking controls–only meQTL models), interaction (i.e., variant-by-smoking status) effects, and joint 2DF tests. Details of each model are described in the Methods and Supplemental Methods. Abbreviations: AA = African ancestry; aai = age of initiation; cpd = cigarettes per day; EA = European ancestry; LIBD = Lieber Institute for Brain Development; meQTL = methylation quantitative trait locus; NAc = nucleus accumbens; sc = smoking cessation; si = smoking initiation.

**Supplemental Table S7. GSCAN – QTL Lookup Summary.** Results included in Supplemental Tables S5 and S6 are summarized for each GSCAN smoking phenotype and for eQTLs and meQTLs separately. Significance thresholds used in tabulation of values are based on the number of overlapping, linkage disequilibrium pruned GSCAN significant variants with each QTL map (see *Supplemental Methods*).

**Supplemental Table S8. NAc meQTL + GSCAN GWAS Colocalization – Summary statistics for colocalization analysis between NAc meQTLs and GSCAN GWAS smoking traits.** Each row corresponds to a colocalization test between variant–CpG associations for a given CpG from *cis*-meQTL mapping and GSCAN GWAS association statistics for smoking initiation, age of initiation, cigarettes per day, or smoking cessation. Because the colocalization test regions may not fully overlap with the predefined GSCAN loci, the boundaries for both the test regions and the GSCAN loci are reported. CpGs corresponding to the meQTLs tested for colocalization were annotated as described in the Supplemental Methods. CpGs for which there is at least one significant variant–CpG association (adjusted p-value$_{2DF}$<0.05) are indicated. The coloc hypotheses are as follows: H0 = no genetic association with either trait; H1 = genetic association with the CpG but not the GSCAN trait; H2 = genetic association with the GSCAN trait but not the CpG; H3 = independent genetic associations with the CpG and GSCAN trait; H4 = shared genetic association with the CpG and GSCAN trait.

**Supplemental Table S9. NAc meQTL + GSCAN GWAS Colocalization – Summary statistics for colocalization analysis between NAc eQTLs and GSCAN GWAS smoking traits.** Each row corresponds to a colocalization test between variant–gene associations for a given gene from *cis*-eQTL mapping and GSCAN GWAS association statistics for smoking initiation, age of initiation, cigarettes per day, or smoking cessation. Because the colocalization test regions may not fully overlap with the predefined GSCAN loci, the boundaries for both the test regions and the GSCAN loci are reported. Genes corresponding to the eQTLs tested for colocalization were annotated as described in the Supplemental Methods**.** Genes for which there is at least one significant variant–gene association (adjusted p-value$_{2DF}$ <0.05) are indicated. The coloc hypotheses are as follows: H0 = no genetic association with either trait; H1 = genetic association with the gene but not the GSCAN trait; H2 = genetic association with the GSCAN trait but not the gene; H3 = independent genetic associations with the gene and GSCAN trait; H4 = shared genetic association with the gene and GSCAN trait.

References


1. Markunas, C.A. *et al.* Genome-wide DNA methylation differences in nucleus accumbens of smokers vs. nonsmokers. *Neuropsychopharmacology* **46**, 554-560 (2021).
2. BrainSeq, A.H.B.G.C. BrainSeq: Neurogenomics to Drive Novel Target Discovery for Neuropsychiatric Disorders. *Neuron* **88**, 1078-1083 (2015).
3. Avila-Tang, E. *et al.* Assessing secondhand smoke using biological markers. *Tob Control* **22**, 164-71 (2013).
4. Anderson, C.A. *et al.* Data quality control in genetic case-control association studies. *Nat Protoc* **5**, 1564-73 (2010).
5. Delaneau, O., Howie, B., Cox, A.J., Zagury, J.F. & Marchini, J. Haplotype estimation using sequencing reads. *Am J Hum Genet* **93**, 687-96 (2013).
6. Marchini, J., Howie, B., Myers, S., McVean, G. & Donnelly, P. A new multipoint method for genome-wide association studies by imputation of genotypes. *Nat Genet* **39**, 906-13 (2007).
7. Numata, S. *et al.* DNA methylation signatures in development and aging of the human prefrontal cortex. *Am J Hum Genet* **90**, 260-72 (2012).
8. Jaffe, A.E. *et al.* Mapping DNA methylation across development, genotype and schizophrenia in the human frontal cortex. *Nat Neurosci* **19**, 40-7 (2016).
9. Aryee, M.J. *et al.* Minfi: a flexible and comprehensive Bioconductor package for the analysis of Infinium DNA methylation microarrays. *Bioinformatics* **30**, 1363-9 (2014).
10. Jaffe, A.E. *et al.* Developmental regulation of human cortex transcription and its clinical relevance at single base resolution. *Nat Neurosci* **18**, 154-161 (2015).
11. Houseman, E.A. *et al.* DNA methylation arrays as surrogate measures of cell mixture distribution. *BMC Bioinformatics* **13**, 86 (2012).
12. Jaffe, A.E. *et al.* Developmental and genetic regulation of the human cortex transcriptome illuminate schizophrenia pathogenesis. *Nat Neurosci* **21**, 1117-1125 (2018).
13. Bolger, A.M., Lohse, M. & Usadel, B. Trimmomatic: a flexible trimmer for Illumina sequence data. *Bioinformatics* **30**, 2114-20 (2014).
14. Patro, R., Duggal, G., Love, M.I., Irizarry, R.A. & Kingsford, C. Salmon provides fast and bias-aware quantification of transcript expression. *Nat Methods* **14**, 417-419 (2017).
15. Srivastava, A. *et al.* Alignment and mapping methodology influence transcript abundance estimation. *Genome Biol* **21**, 239 (2020).
16. Love, M.I., Hogenesch, J.B. & Irizarry, R.A. Modeling of RNA-seq fragment sequence bias reduces systematic errors in transcript abundance estimation. *Nat Biotechnol* **34**, 1287-1291 (2016).
17. Soneson, C., Love, M.I. & Robinson, M.D. Differential analyses for RNA-seq: transcript-level estimates improve gene-level inferences. *F1000Res* **4**, 1521 (2015).
18. Kim, D., Paggi, J.M., Park, C., Bennett, C. & Salzberg, S.L. Graph-based genome alignment and genotyping with HISAT2 and HISAT-genotype. *Nat Biotechnol* **37**, 907-915 (2019).
19. Ewels, P., Magnusson, M., Lundin, S. & Kaller, M. MultiQC: summarize analysis results for multiple tools and samples in a single report. *Bioinformatics* **32**, 3047-8 (2016).
20. Danecek, P. *et al.* Twelve years of SAMtools and BCFtools. *Gigascience* **10**(2021).
21. Aschard, H., Hancock, D.B., London, S.J. & Kraft, P. Genome-wide meta-analysis of joint tests for genetic and gene-environment interaction effects. *Hum Hered* **70**, 292-300 (2010).
22. Shabalin, A.A. Matrix eQTL: ultra fast eQTL analysis via large matrix operations. *Bioinformatics* **28**, 1353-8 (2012).
23. Love, M.I., Huber, W. & Anders, S. Moderated estimation of fold change and dispersion for RNA-seq data with DESeq2. *Genome Biol* **15**, 550 (2014).

24. Stegle, O., Parts, L., Piipari, M., Winn, J. & Durbin, R. Using probabilistic estimation of expression residuals (PEER) to obtain increased power and interpretability of gene expression analyses. *Nat Protoc* **7**, 500-7 (2012).
25. Huang, Q.Q., Ritchie, S.C., Brozynska, M. & Inouye, M. Power, false discovery rate and Winner's Curse in eQTL studies. *Nucleic Acids Res* **46**, e133 (2018).
26. Davis, J.R. *et al.* An Efficient Multiple-Testing Adjustment for eQTL Studies that Accounts for Linkage Disequilibrium between Variants. *Am J Hum Genet* **98**, 216-24 (2016).
27. Subramanian, A. *et al.* Gene set enrichment analysis: a knowledge-based approach for interpreting genome-wide expression profiles. *Proc Natl Acad Sci U S A* **102**, 15545-50 (2005).
28. Liu, M. *et al.* Association studies of up to 1.2 million individuals yield new insights into the genetic etiology of tobacco and alcohol use. *Nat Genet* **51**, 237-244 (2019).
29. Chang, C.C. *et al.* Second-generation PLINK: rising to the challenge of larger and richer datasets. *Gigascience* **4**, 7 (2015).
30. Pers, T.H., Timshel, P. & Hirschhorn, J.N. SNPsnap: a Web-based tool for identification and annotation of matched SNPs. *Bioinformatics* **31**, 418-20 (2015).
31. Foley, C.N. *et al.* A fast and efficient colocalization algorithm for identifying shared genetic risk factors across multiple traits. *Nat Commun* **12**, 764 (2021).
32. Oscanoa, J. *et al.* SNPnexus: a web server for functional annotation of human genome sequence variation (2020 update). *Nucleic Acids Res* **48**, W185-W192 (2020).
